# Emulator-based Bayesian calibration of the CISNET colorectal cancer models

**DOI:** 10.1101/2023.02.27.23286525

**Authors:** Carlos Pineda-Antunez, Claudia Seguin, Luuk A van Duuren, Amy B. Knudsen, Barak Davidi, Pedro Nascimento de Lima, Carolyn Rutter, Karen M. Kuntz, Iris Lansdorp-Vogelaar, Nicholson Collier, Jonathan Ozik, Fernando Alarid-Escudero

**Author notes:** **Corresponding Author:** Fernando Alarid-Escudero, Ph.D., 615 Crothers Way, #117, Encina Commons, MC 6019, Stanford, CA 94305. e-mail address.

## Abstract

**Purpose:** To calibrate Cancer Intervention and Surveillance Modeling Network (CISNET) ’s SimCRC, MISCAN-Colon, and CRC-SPIN simulation models of the natural history colorectal cancer (CRC) with an emulator-based Bayesian algorithm and internally validate the model-predicted outcomes to calibration targets.

**Methods:** We used Latin hypercube sampling to sample up to 50,000 parameter sets for each CISNET-CRC model and generated the corresponding outputs. We trained multilayer perceptron artificial neural networks (ANN) as emulators using the input and output samples for each CISNET-CRC model. We selected ANN structures with corresponding hyperparameters (i.e., number of hidden layers, nodes, activation functions, epochs, and optimizer) that minimize the predicted mean square error on the validation sample. We implemented the ANN emulators in a probabilistic programming language and calibrated the input parameters with Hamiltonian Monte Carlo-based algorithms to obtain the joint posterior distributions of the CISNET-CRC models’ parameters. We internally validated each calibrated emulator by comparing the model-predicted posterior outputs against the calibration targets.

**Results:** The optimal ANN for SimCRC had four hidden layers and 360 hidden nodes, MISCAN-Colon had 4 hidden layers and 114 hidden nodes, and CRC-SPIN had one hidden layer and 140 hidden nodes. The total time for training and calibrating the emulators was 7.3, 4.0, and 0.66 hours for SimCRC, MISCAN-Colon, and CRC-SPIN, respectively. The mean of the model-predicted outputs fell within the 95% confidence intervals of the calibration targets in 98 of 110 for SimCRC, 65 of 93 for MISCAN, and 31 of 41 targets for CRC-SPIN.

**Conclusions:** Using ANN emulators is a practical solution to reduce the computational burden and complexity for Bayesian calibration of individual-level simulation models used for policy analysis, like the CISNET CRC models. In this work, we present a step-by-step guide to constructing emulators for calibrating three realistic CRC individual-level models using a Bayesian approach.

## Introduction

Individual-based simulation models are often used to evaluate health policies that aim to reduce the impact of diseases (1). For many of these models, data to directly inform the parameters are lacking and often estimated through calibration. Ideally, the uncertainty in model parameters should be accurately quantified during the calibration process so that the ultimate model predictions also reflect this uncertainty (2). Therefore, the calibration process aims to obtain a joint posterior distribution of the calibrated parameters that produce model outputs consistent with the calibration targets (observed data arising from a clinical or epidemiological system) and their uncertainty (3). Health policy models are often complex, with a relatively large number of parameters, making calibration challenging. (4).

Bayesian methods are suitable for calibrating health decision models because they provide a framework to estimate parameter uncertainty (5), instead of just obtaining point estimates, which is consistent with current modeling guidelines and recommendations (6,7). In addition, under a Bayesian framework, researchers could include prior knowledge or beliefs about the parameters in the model calibration process, which is especially valuable when limited data is available. However, there are many challenges involved in Bayesian calibration. The computing time for calibrating complex models using Bayesian algorithms, such as the incremental mixture importance sampling (IMIS), (8) is significantly higher than direct search methods. Bayesian algorithms also demand high computational power, requiring running the model thousands or even millions of times to achieve convergence (9). For example, IMIS requires an initial draw from the prior distribution of several thousands of samples; for each of them, a sampling weight is computed by evaluating the likelihood using the simulation model, a new draw of thousands of samples is generated using the sampling weights and iteratively continues to repeat this process until a maximum number of iterations is reached, or a specific effective sample size is achieved. All this process requires evaluating the model several tenths or hundreds of thousands of times (10), which can be highly computationally demanding.

One approach to reducing the computational burden uses an emulator or metamodel that serves as a surrogate of the original complex simulation model (often referred to as a simulator) by mapping the relationship between the inputs and outputs of the simulator (11,12). An emulator is a proxy model that is less complex and faster than a simulator, can replace a simulator to predict outcomes, and is often used in other model-based analyses, such as calibration procedures (8), policy optimization (11,13), cost-effectiveness analysis (14), extrapolating findings to other settings (15) and probabilistic analysis (16). Using emulators for Bayesian calibration can reduce the computational time to obtain a sample from the model parameters posterior distribution, which has already been shown to work with a considerable reduction of time (8,17).

This work aims to quantify the uncertainty of parameters of three microsimulation models of the natural history of colorectal cancer (CRC) using Bayesian Calibration with Artificial Neural Networks (BayCANN), an emulator-based algorithm (8) and validate the model-predicted outcomes to calibration targets.

## Methods

We used three models from the Cancer Intervention and Surveillance Modeling Network (CISNET) Colorectal Cancer (CRC) Working Group - Simulation Model of Colorectal Cancer (SimCRC), Microsimulation Screening Analysis (MISCAN-Colon), and Colorectal Cancer Simulated Population model for Incidence and Natural history (CRC-SPIN) –focused on the long-term objective of reducing the burden of CRC. These microsimulation models describe the natural history of CRC using different underlying structures(18). They have a long history of use in comparative modeling analyses to provide the information needed to address key policy questions and prioritize future research and have been used to evaluate various health policies and clinical strategies, such as screening and surveillance of CRC programs in the US (19,20).

The CISNET CRC models can be used to generate counterfactual scenarios in which one or more factors are changed based on a simulated policy to calculate the change in expected benefits, harms, and costs in the target population. For each simulated individual, age-, and sex-specific risks affect the number and age of onset of adenomas, i.e., precancerous tumors. Each adenoma inside an individual can have a different size and location in the large intestine; some can transition to cancer. The simulation of the natural history of CRC allows us to evaluate screening policies that aim to interrupt the adenoma-carcinoma sequence. The targets included in the models are outcomes related to the natural history of CRC, and parameters are part of the functions that simulate the different events of the natural history of CRC

Under the BayCANN algorithm, we construct ANNs as emulators replacing the CISNET CRC models for Bayesian calibration. In the following sections, we describe six steps to implement BayCANN. An illustration of the BayCANN process is shown in Figure 1.

**Figure 1.**
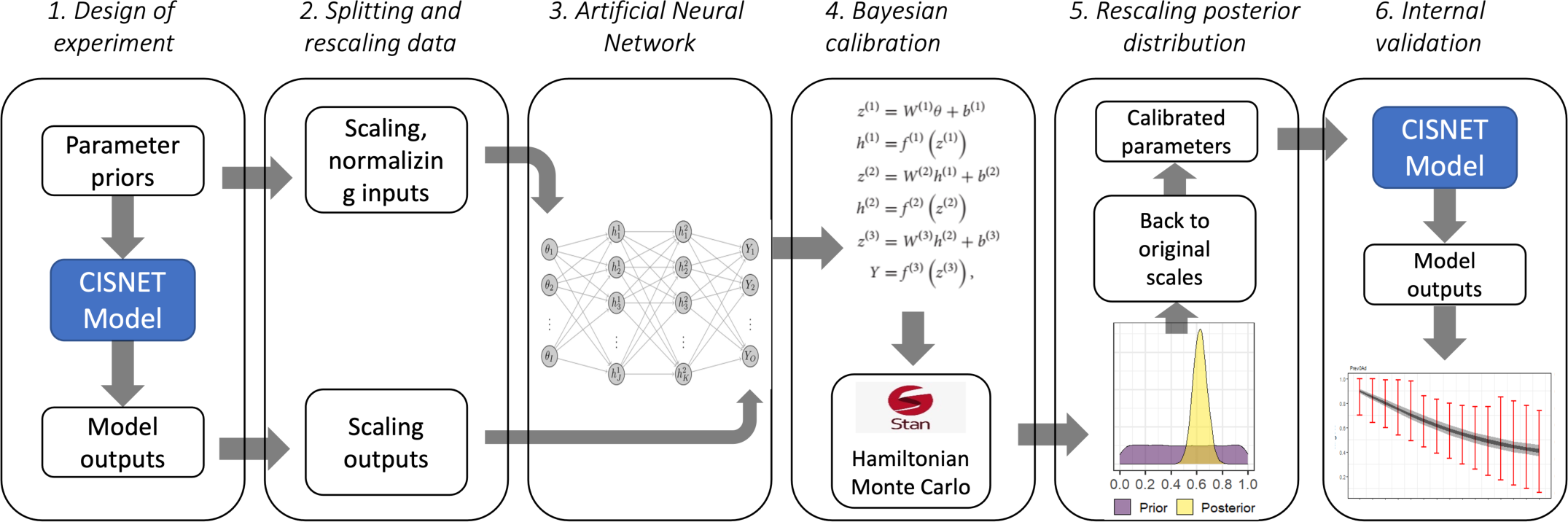
Diagram of BayCANN implementation.

### Design of experiment

The first step in designing an emulator is obtaining a set of inputs and their corresponding output values from the original model using a design of simulation experiment (21). These input-output pairs are required to map the underlying structure of the models. We used Latin Hypercube Sampling (LHS) to generate model parameter inputs that efficiently sample the parameter space. We designed the LHS to ensure that the parameter values sampled would lead to model outputs containing the mean of the calibration targets. This means that the model-predicted outputs obtained from the LHS parameter space can produce, or cover, the target’s mean and uncertainty measure (e.g., 95% confidence interval). We denote this exercise as a “coverage analysis”. A low performance in the coverage analysis, meaning the data used to train the emulator does not contain values consistent with the calibration targets (e.g., producing values consistently lower or higher than the targets),will prevent the emulator from producing posterior distributions that will make the model fit the targets.

To obtain the model outputs, CRC-SPIN and MISCAN-Colon run simulations with different population sizes for each calibration target, based on previous analysis showing these sample sizes obtained stable outcomes. The population size ranged from 50 thousand to 10 million population. SimCRC used a unique population of 10 million. In this study, as in any model calibration, the dimensionality of the model outputs is equal to the number of calibration targets. CRCSPIN and MISCAN used the Theta supercomputer from Argonne Leadership Computing Facility where each computing node has 64-core, 1.3-GHz Intel Xeon Phi 7230 (22). SimCRC performed the analyses using distributive computing on a cluster of machines that together have 36 cores. The majority of processes were performed on a machine with an 18-core

### 3.0 GHz Intel i9-10980XE processor

#### Splitting samples and rescaling inputs and outputs

For the second step, we prepare data for the emulator training. First, we split the LHS data into training and testing sets using 80% and 20% of the data, respectively, to reduce overfitting. Second, we scaled the inputs and outputs from -1 to 1 to have all the data on the same scale because different magnitudes between and within inputs and outputs could reduce the performance and efficiency of training the ANN (23). We also test if normalizing the model inputs, meaning that we adjusted the prior distributions to make them normally distributed, improves the prediction performance.

#### Constructing the Artificial Neural Networks

In the third step, we build the emulator using a Multilayer Perceptron (MLP), which consists of one input layer, one or more hidden layers, and one output layer, each consisting of nodes that are all connected between layers, often referred to as a fully connected class of ANN (23) The input layer takes in the different values for each model input parameter and has as many nodes as the number of the simulator input parameters to be calibrated. The output layer contains as many nodes as the number of the simulator model outputs to be used to compare to the calibration targets. The number of hidden layers and nodes are considered hyperparameters and are determined based on the fit of the ANN to the model outcomes. We used *`Keras`* (version 2.8.0) (24) and *`TensorFlow`* (version 2.8.0) (25) packages in R to build and train the ANN emulators.

To find the optimal hyperparameters, we performed a grid search using a full factorial design by varying the number of hidden layers between one and five and the number of hidden nodes per hidden layer starting from the number of model outputs up to 300 more with increments of 20. Because each CISNET CRC model has a different number of input parameters and outcomes, we applied the grid search for each model. Once we define an optimal ANN emulator structure, we tuned other hyperparameters. These include the selection of activation functions (sigmoidal, hyperbolic tangent, relu), batch size (500, 1000, 2000), and the optimizer (Adam, gradient descent), to improve the performance prediction of the ANN emulator. For each CISNET CRC model, we selected the structure that minimizes the mean square error (MSE) on the validation set.

Equation (1) describes the emulator equation for an MLP ANN emulator for the three CISNET-CRC models.

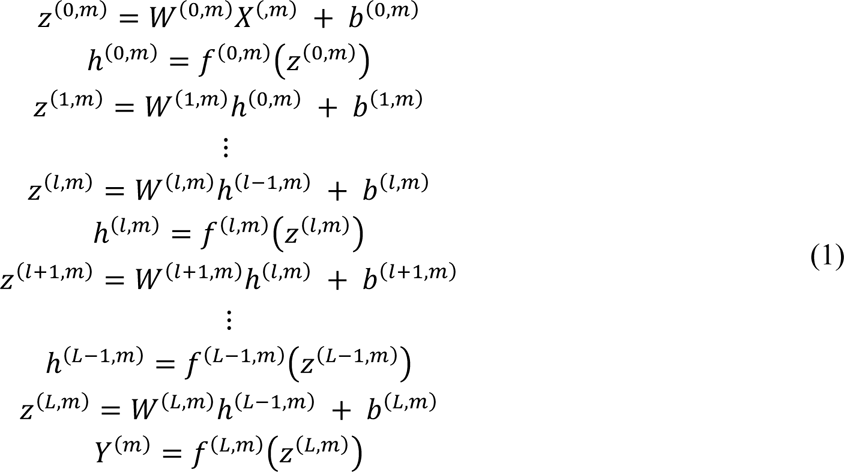

Where *m* indicates the model (SimCRC, MISCAN-Colon, CRCSPIN); *l* = 0, …, *L*, indicates the layer (*l* = 0 is the input layer, *l* = *L* is the output layer, and 0 < *l* < *L* represent a hidden layer); *Z*^(*l,m*)^ is the weighted sum of inputs to the nodes in layer *l* and model *m*; ***X***^(*m*)^ = \**X*_1,*m*_, *X*_2,*m*_, …, *X*_1,*x*_ + is the vector of parameters (or inputs) for model *m*; *W*^(*l,m*)^ is a matrix of weights; *b*^(*l,m*)^ is a vector of biases, *h*^(*l,m*)^is the value of the activation function, *f*^(*l,m*)^, evaluated in *Z*^(*l,m*)^, which is the input for the nodes in the next layer. An activation function on layer *l* aggregates a non-linear relationship between layers (*l* − 1) and *l*. Finally, *Y*^(*m*)^ = \**Y*_&,$_, *Y*_+,$_, …, *Y*_.,$_ + is the vector of predicted-emulator outcomes for model *m* and total outcomes *O*.

We evaluated the goodness of fit of the emulators in reproducing the CISNET-CRC model-predicted outcomes by estimating the average R-square between each pair of emulators and simulator output in the LHS test sample(26).

#### Bayesian calibration

In this study, the parameters set corresponds to a vector model parameters that need to be estimated via calibration to make the model outputs consistent with calibration targets. Let 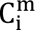 denote the i-th element (e.g., an age-specific estimate) for the calibration target set C (e.g., CRC incidence or adenoma prevalence, etc.) for model m. The process of model calibration is an estimation problem where we seek to estimate the parameters for model m, θ^0^, by using a summary measure of the discrepancy between the corresponding model output, 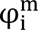, and 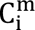. For this analysis, we use the likelihood function as the summary measure assuming the calibration targets 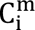 are normal deviations from the model outputs 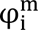 with standard deviation 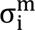.

That is,

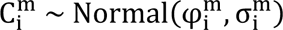

Therefore, the likelihood function is,

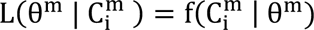

Where 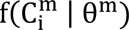 denotes the normal density function for the target 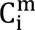

For the purposes of doing the Bayesian calibration with the ANN emulator, 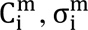, and 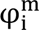 represent the rescaled values of the targets, standard deviations, and model outputs, respectively, based on the transformation functions described in section *Splitting samples and rescaling inputs and outputs*.

In the fourth step, we implemented the selected ANN structure for each CRC model by coding the emulator equation in Stan (27). In this step, the emulators replaced the CRC models within the Bayesian calibration algorithm. Using four Hamiltonian Monte Carlo (HMC) chains, we ran the algorithm for 100,000 samples to obtain a sample of 1,000 draws from the posterior distribution. We verified the mixing and convergence of the chains using the R-hat convergence diagnostic. If chains have not mixed well, the R-hat is greater than 1.2. We used the HMC algorithm in Stan to sample from the posterior distributions according to a target probability distribution. The HMC requires less computational time than Metropolis or Gibbs sampling, because it requires fewer samples to converge to the posterior distribution. This advantage becomes particularly relevant as models increase in complexity, allowing HMC to significantly outperform alternative algorithms (28).

#### Rescaling posterior distribution to original input parameter scale

In the fifth step, after obtaining the posterior distributions using the emulator, we transformed the inputs back to the original scale to use them in the corresponding CISNET model to validate that the model outputs fit their related calibration targets.

#### Internal validation

Lastly, in the sixth step, we internally validated each calibrated simulator using a Bayesian posterior predictive check by propagating the uncertainty of the parameter’s posterior distributions on model-predicted outputs and comparing them to the calibration target means and 95% confidence intervals. For each CISNET-CRC model, we used BayCANN to draw a sample of 1,000 calibrated parameter sets from their posterior distribution and ran the models to produce their corresponding outputs.

The data and code to perform BayCANN for the three CRC CISNET models are available at the GitHub repository: https://github.com/NCI-CISNET-Colorectal/baycann_cisnet_crc.

## Results

### Inputs and outputs from the simulation models

Using an LHS design, we sampled 50,000, 37,000, and 16,900 parameter sets and generated corresponding outputs for the SimCRC, MISCAN-Colon, and CRC-SPIN models, respectively. Each model generated a different LHS size given the number of parameters to be calibrated and computer capacity, ensuring a minimal sample size based on a commonly used rule of *sample size* ≥ 10 ∗ *I* ∗ *O* where *I* is the number of model parameters (inputs) and O is the number of targets (outputs) (29). The average time the models took for each run was 5.6, 5.0, and 3.8 minutes/core for the SimCRC, MISCAN-Colon, and CRC-SPIN models, respectively.

The calibrated models varied in terms of the coverage analysis for the calibration targets means. SimCRC covered 100% of the targets, MISCAN-Colon covered 94% and CRCSPIN covered 85%. The targets’ coverage for each CISNET-CRC model is shown in Figure S1 in the supplementary material.

### ANN emulators of CISNET-CRC models

The number of hidden layers and nodes from the ANN structure with the lowest MSE varied across models. SimCRC’s ANN had four hidden layers with 360 hidden nodes, MISCAN-Colon’s ANN had four hidden layers with 114 hidden nodes, and CRC-SPIN’s ANN had one hidden layer with 140 hidden nodes. The computer used for emulator training had 8 cores and 32 GB RAM. The duration of training varied for different models, with 39, 26, and 10 minutes taken for SimCRC, MISCAN-Colon, and CRC-SPIN, respectively, as shown in Table 1. The training time was influenced by the structure of the ANNs as well as the number of samples utilized in each LHS design. We selected the input transformation that yielded the best prediction performance, including normalization and rescaling from -1 to 1. All model inputs benefited from scaling them between -1 and 1. Normalizing the inputs improved the prediction performance only for CRCSPIN. We did not observe an additional benefit for the other two models. The more hidden nodes or layers and the more samples to train, the longer it took to train them.

**Table 1.** Transformations, structure, training, and calibration time of the ANN emulator selected for each CISNET model.

| Model | LHS samples and time (hours)* | Required transformation | Best out-of-sample ANN Structure | Training time | Calibration time |
| --- | --- | --- | --- | --- | --- |
| **SimCRC**<br>*30 parameters*<br>*110 targets* | 50,000 samples<br>72 hours | - Scale inputs (-1 to 1)<br>- Scale outputs (-1 to 1) | - 4 hidden layers<br>- 360 hidden nodes<br>- Activation function: Hyperbolic tangent | 39 min | 6.7 hours |
| **MISCAN-Colon**<br>*37 parameters*<br>*93 targets* | 37,000 samples<br>48 hours | - Scale inputs (-1 to 1)<br>- Scale outputs (-1 to 1) | - 4 hidden layers<br>- 114 hidden nodes<br>- Activation function: Hyperbolic tangent | 26 min | 3.6 hours |
| **CRC-SPIN**<br>*22 parameters*<br>*41 targets* | 18,000 samples<br>16 hours | - Normalize inputs $\sim N(0,1)$<br>- Scale outputs (-1 to 1) | - 1 hidden layer<br>- 140 hidden nodes<br>- Activation function: Sigmoid | 10 min | 0.5 hours |

\*LHS time was estimated assuming 64 cores for the three models

We obtained goodness of fit in terms of the average R-squared of 0.987, 0.997, and 0.899 for SimCRC, MISCAN-Colon, and CRC-SPIN emulators, respectively. Figure 2 shows the prediction performance of the three emulators, where each point is the pair of CISNET-CRC model (X-axis) and emulator prediction (Y-axis) of a sample of CRC incidence targets for a given parameter set in the LHS, and the 45° line means a perfect prediction. For this sample of incidence outputs, the MISCAN-Colon emulator had the best performance of all the emulators. Overall, most of the ANN outputs had a good prediction performance across all the range of the model outputs covered in the LHS. This range of model outputs encompasses the target values as well. In supplementary material S6, we show the model outputs’ prediction performance.

**Figure 2.**
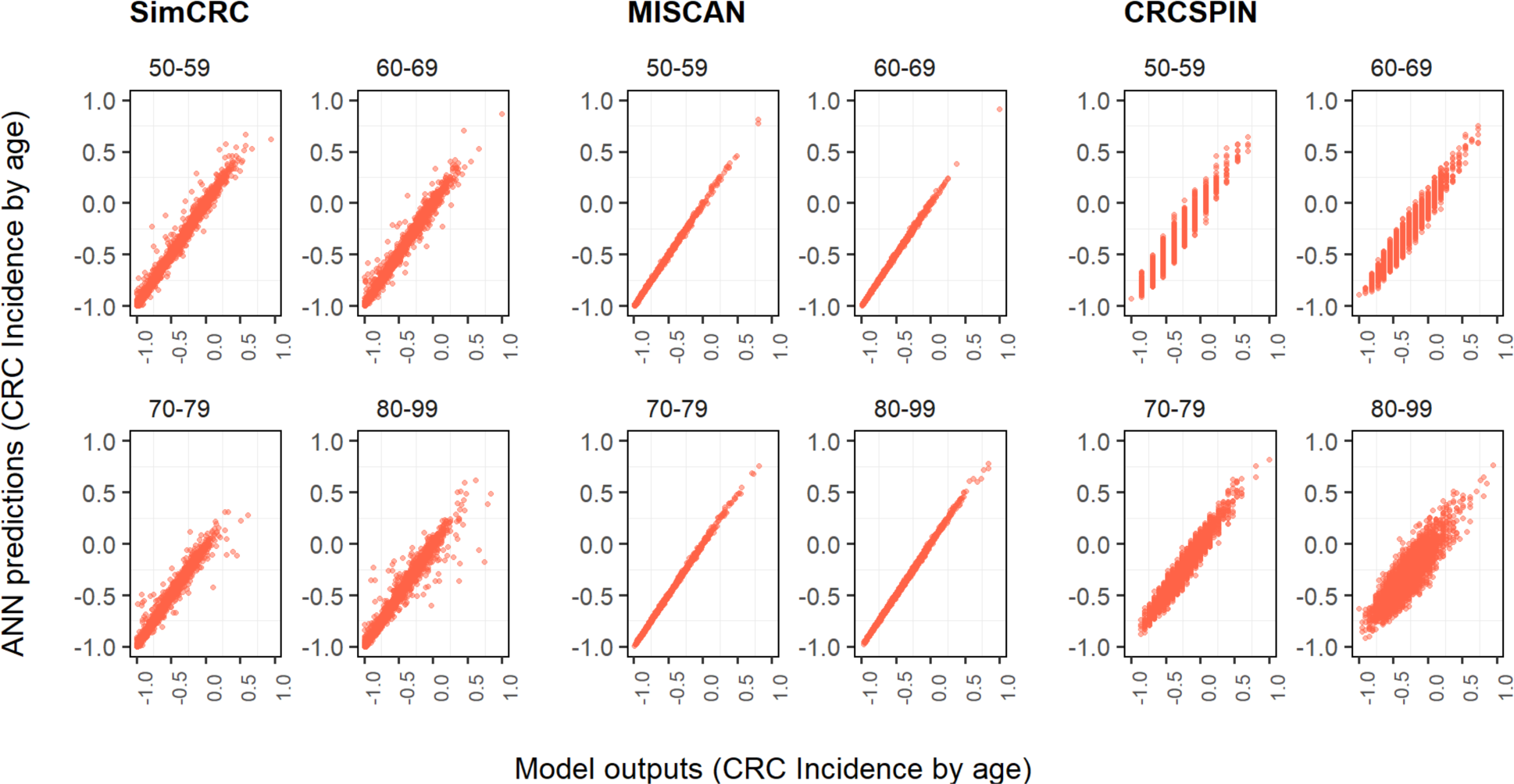
ANN predictions. (y-axis) vs. model outputs (x-axis) for four age groups of CRC incidence targets Note: CRCSPIN generates discretized values for incidence, and the ANN generates continuous values. Therefore, different continuous prediction values are made for one discrete value from CRCSPIN”

The time each emulator spent to estimate outputs given a set of input parameters was 0.079, 0.025, and 0.031 milliseconds for SimCRC, MISCAN-Colon, and CRCSPIN, respectively. These times represent a reduction of up to eight orders of magnitude of the time used by the CRC-CISNET models to produce the same outputs.

### Bayesian calibration

We used the final ANN emulators and implemented them in Stan to obtain the calibrated posterior distributions of the parameters using four independent chains. For all three CISNET-CRC emulators, we obtained good convergence and mixing for the four chains of the posterior distribution with an R-hat less than 1.003 for all parameters of the three CISNET-CRC models. Values of the R-hat smaller than 1.2 have been set as thresholds for satisfactory convergence (30). Additional diagnostics of the Bayesian calibration, such as R-hat (for convergence) and Effective Sample Size (for efficiency), are shown in Figure S2 in the supplementary material. Most of the parameters’ chain correlation converged to zero after the first iterations, which is also an indicator of good mixing on MCMC chains. The time required to calibrate the emulators differed by model, with longer times for models with more inputs and outputs. The times for calibrating the emulator were 6.7, 3.6, and 0.5 hours for SimCRC, MISCAN-Colon, and CRC-SPIN, respectively.

### Calibrated posterior distributions

After the calibration, the posterior distribution of the calibrated parameters reduced the uncertainty compared to the prior distributions. The level of reduction varied between the models. For example, Figure 3 illustrates that the *hazard_mean_uk* parameter in the MISCAN-Colon model showed a more pronounced uncertainty reduction than *pSxDetS1_D* and *growth.rectum.beta1* parameters in SimCRC and CRC-SPIN parameters, respectively. In general, most of the posterior distributions shrunk compared to their priors. The prior and posterior marginal distributions for all model parameters are shown in Figure S3 in the supplementary material.

**Figure 3.**
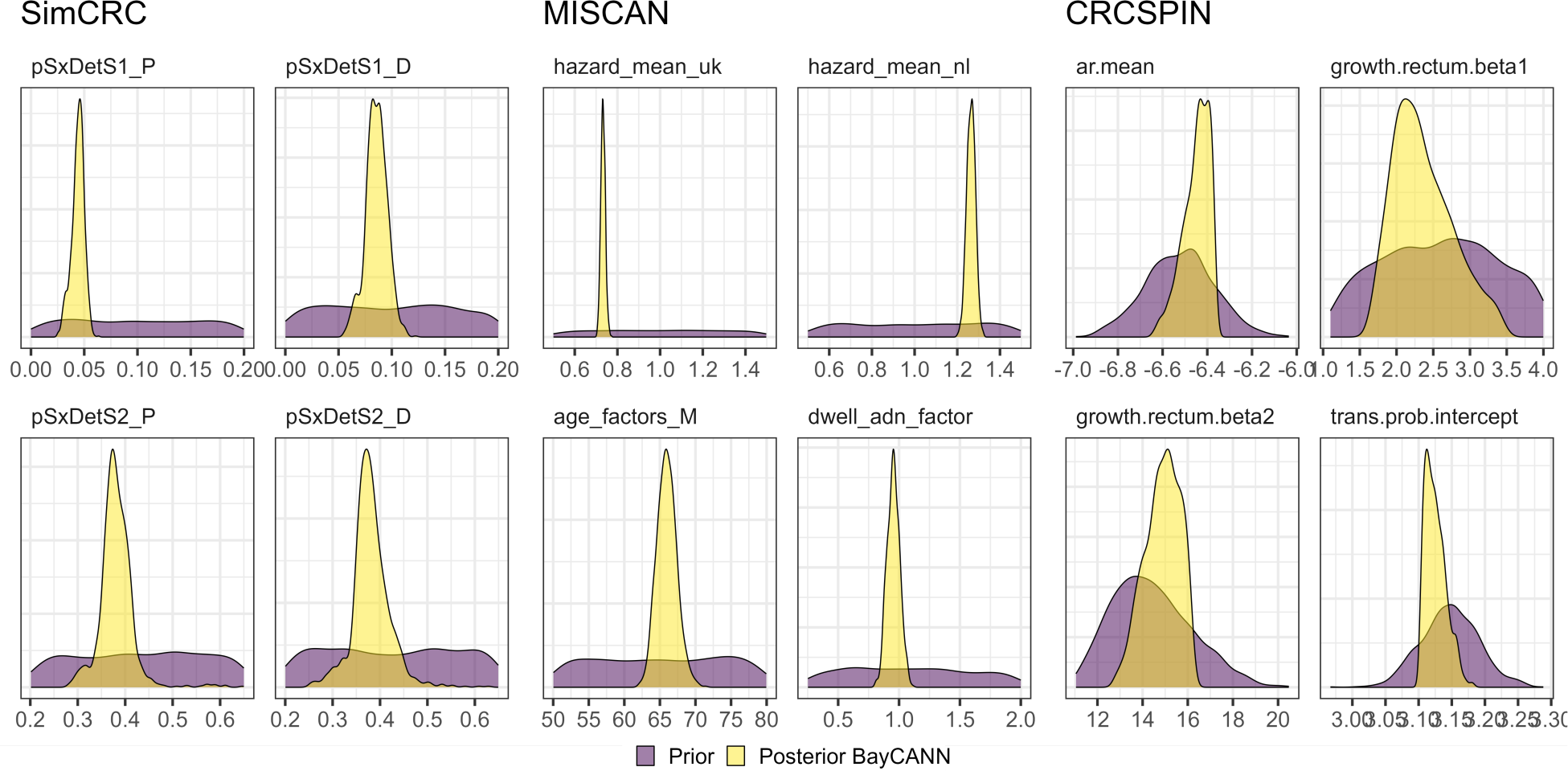
Prior and posterior marginal distributions of calibrated parameters for the three CISNET-CRC models.

The posterior distribution for MISCAN-Colon had the pairs of parameters with the highest correlation values reaching 0.99, followed by SimCRC with 0.93 and 0.74 for CRC-SPIN. The joint posterior distributions of all pairwise parameters for the three CISNET-CRC models are shown in Figure S4 in the supplementary material. Highly correlated parameters are often found in these types of simulation models with a high number of parameters (2)

### Internal validation

The calibrated models showed good internal validation. Specifically, the interquartile range of the predictions of adenoma prevalence was within the target interval for all age groups in the three CISNET-CRC models. Moreover, the CRC incidence predictions were within the target interval for most age groups, as depicted in Figure 4. Overall, the mean predicted outcomes that fell within the 95% confidence intervals of the calibration targets for SimCRC were 98 of 110, 65 of 93 for MISCAN, and 31 of 41 for CRC-SPIN (see Figure S5 in the supplementary material).

**Figure 4.**
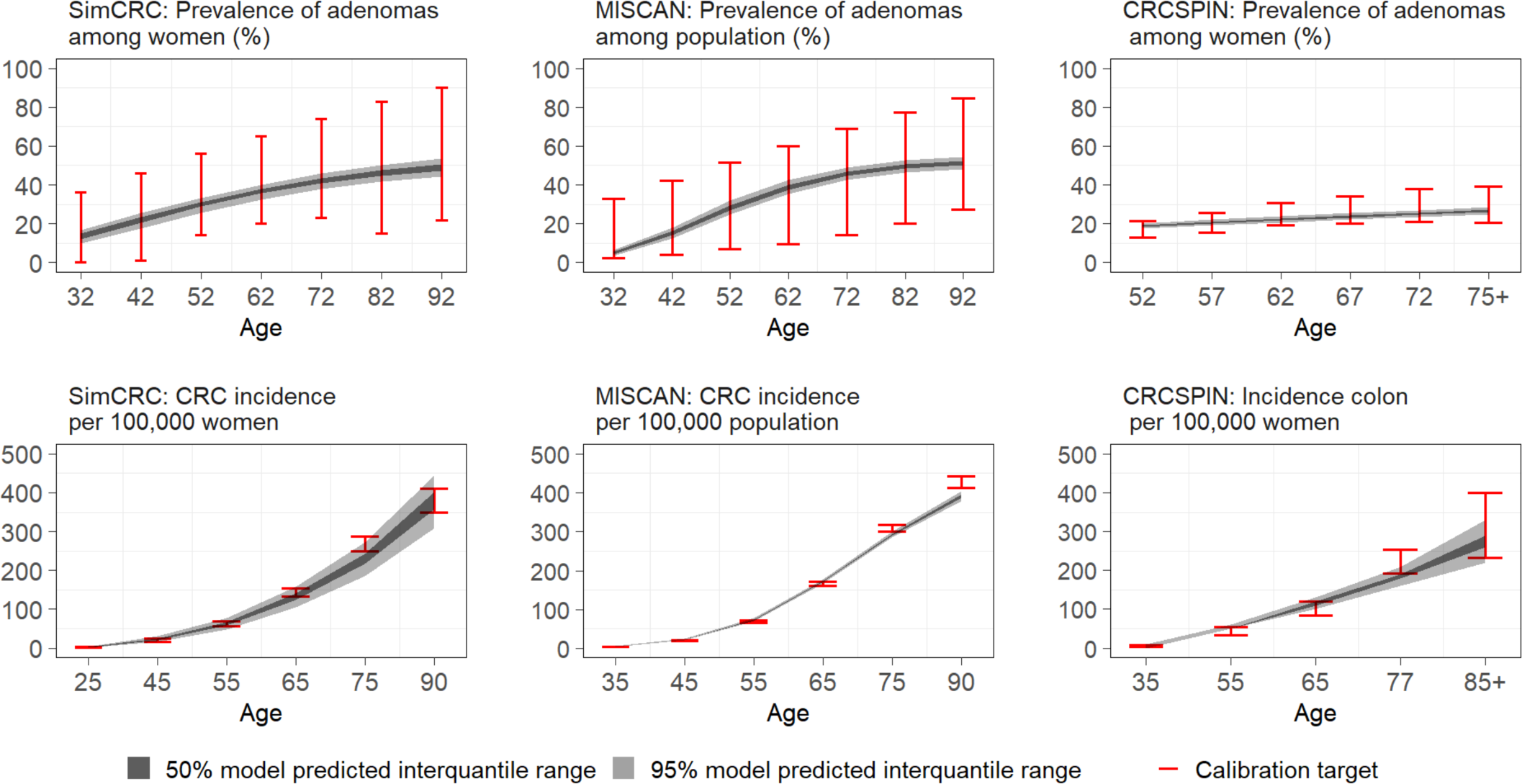
Validation for prevalence and incidence targets of SimCRC, MISCAN-Colon and CRC-SPIN models.

## Discussion

In this manuscript, we used ANN emulators of the CISNET-CRC models as a practical solution to reduce the computational burden of calibration and efficiently calibrate these models under a Bayesian framework. Although emulators have been used widely in other science and engineering areas, they have been underused in health decision science. This may be due to a lack of material and guidance on developing and validating them (31). To address these issues, we provided step-by-step guidance on constructing ANN emulators to calibrate three realistic CRC microsimulation models using a Bayesian approach. We showed that BayCANN serves as a flexible framework for building a suitable emulator for CISNET models that avoids the need to try multiple emulator alternatives, such as Gaussian process regression, generalized additive models, symbolic regression, etc., that also can be suitable metamodels but require more trial and error and consequently investing more time.

In a previous study using BayCANN on a CRC simulation model, the authors showed that BayCANN was faster and more accurate in recovering the true parameter values than calibrating the simulation model directly with the incremental mixture importance sampling (IMIS) algorithm, which is often used for Bayesian calibration (8). Regarding the three CRC-CISNET models, only CRCSPIN has been previously calibrated using Bayesian methods under a likelihood-free approach with the incremental mixture approximate Bayesian computation (IMABC) algorithm (32). The posterior distribution for CRCSPIN obtained from BayCANN produced similar outputs to those obtained from IMABC, but the latter required 2,352 node hours (32). MISCAN-Colon and SimCRC were previously calibrated but using direct-search approaches, such as Nelder-Mead and simulated annealing, which only generated single best-fitting parameter sets.

As in previous studies, our emulators proved to be an efficient alternative for conducting analysis based on complex microsimulation models and enabling computationally intensive processes such as Bayesian calibrations (11,33). The emulator-based Bayesian calibration conducted in this work provides a fast solution to calibrate multitarget models and obtain uncertainty of the model parameters. Using emulators instead of the original simulation models allowed us to perform significantly more iterations in the calibration process, which helped to improve the results. Our results showed a good calibration performance for the three CISNET-CRC models, generating outputs within targets’ uncertainty for most of their outcomes (>70%).

We found that as the complexity of the simulation model structure in terms of the number of input parameters (i.e., ANN inputs) and target outcomes (i.e., ANN outputs) increases, the more layers the ANN emulator requires, resulting in a higher number of the ANN parameters that need to be estimated. For example, SimCRC with 30 parameters and 110 targets required an ANN emulator with 4 layers with 360 nodes each and a total of 571,070 ANN parameters, MISCAN with 37 input parameters and 96 target outcomes required an ANN emulator with 4 layers with 114 nodes each and a total of 67,467 ANN parameters and CRCSPIN with 22 parameters and, 41 targets required an ANN with one 1 layer and 140 nodes each and a total of 28,741 ANN parameters. Considering the total ANN parameters for each model, the increase in training time did not increase exponentially as a function of the number of ANN parameters need to be estimated (see Figure S7 in Supplementary Material), which is consistent with previous studies (34).

ANNs showed good performance in emulating the CISNET-CRC microsimulation models. Although there are advantages and disadvantages of specific emulator algorithms that may vary across different model applications, our framework for metamodeling using ANN can be employed in applications with a high number of inputs and outputs, which is often the case with simulation models in the field of medical decision making. Although GPs have shown highly accurate predictions in metamodeling studies using small datasets or few inputs and outputs, the time to fit the GP emulator increases exponentially as the number of parameters in the simulator that require calibration increases, eventually becoming an intractable problem (11,16,35). In these cases, neural networks are more suitable because they scale better in terms of dimensionality than GPs and other emulators(36). Furthermore, ANNs are relatively easy to construct in commonly used programming languages and implement in Bayesian software, such as JAGS or Stan, for calibration purposes. In our study, we construct emulators of real-world simulation models used to inform screening and treatment strategies with a high number of input parameters and outputs. For these reasons, ANNs are good candidates as emulators of individual-based models for Bayesian calibration.

Our findings are subject to the limitations of the neural networks models used as emulators. Determining the ANN’s hyperparameter values a priori can be challenging and requires some supervision in the training process. For example, we need to define the number of hidden layers and nodes, which must be sufficiently high to capture the underlying functions but not so high as to cause overfitting, which is not ideal. We split the LHS sample into training and testing sets to prevent overfitting. The testing sets were used to validate the predictions not used in the training process. We limited our search space for ANN hyperparameters that ensured reaching an accuracy of more than 90%. Achieving higher accuracy levels would likely require more computational time and burden, diminishing the benefits of using emulators.

The reduced computational burden does not consider the time required to generate the LHS design, which could be considerable because the original individual-level models are required for this process. However, an LHS is also required for other commonly used Bayesian calibration algorithms, such as IMIS (37), and IMABC (32). The number of LHS observations required to train the emulators depends on the underlying functions of each microsimulation model; we did not explore the optimal number of LHS observations needed for our emulators, so we could potentially reduce this number to decrease the time it takes to generate the LHS without affecting the learning process of the emulators. A possible extension to our approach includes implementing adaptative sampling with emulators to identify the optimal number of LHS observations; this exercise may reduce the overall calibration time (38,39).

BayCANN is a transparent and reproducible algorithm that can facilitate the Bayesian calibration of individual-based models. It can be implemented in commonly used open-source software, such as R or Python (8). Bayesian calibration typically requires many model evaluations; we performed at least 100,000 iterations within the Hamiltonian Monte Carlo algorithm for this work. Using the CISNET models that require, on average, 4 minutes per iteration (using similar computational resources) may become prohibitively expensive.

BayCANN, as with other emulator-based calibration approaches, requires the original simulator only to generate the LHS design and for internal validation. The actual Bayesian calibration is done with the trained emulator using the LHS samples. An advantage of using emulators for model calibration is that it is not required to code the simulator in probabilistic software (e.g., Stan), which represents another time-consuming task to perform the calibration or may even be impossible for likelihood-free models. Another advantage is that we could use likelihood-based Bayesian calibration approaches in situations when this would not be feasible or require simulating a substantially large population to simulate the likelihood, and likelihood-free approaches are often used. BayCANN can be applied to calibrate other computationally intensive individual-level decision models and contribute to the availability of algorithms that facilitate the implementation of Bayesian calibration.

## Conclusion

In this study, we showed that ANNs are suitable emulators of the CISNET CRC models for Bayesian calibration. The use of emulators significantly reduced the time and computational burden for calibration. Analysts wanting to calibrate computationally-expensive simulation models to quantify calibrated parameter uncertainty accurately under a Bayesian framework could benefit from using BayCANN.

## Data Availability

All data produced in the present study are available upon reasonable request to the authors

## Acknowledgments

Financial support for this study was provided by the grant U01-CA253913 from the National Cancer Institute (NCI) as part of the Cancer Intervention and Surveillance Modeling Network (CISNET). The content is solely the responsibility of the authors and does not necessarily represent the official views of the National Institutes of Health. The funding agencies had no role in the design of the study, interpretation of results, or writing of the manuscript. The funding agreement ensured the authors’ independence in designing the study, interpreting the data, writing, and publishing the report. This research was completed with resources provided by the Argonne Leadership Computing Facility (Theta), which is a DOE Office of Science User Facility supported under Contract DE-AC0206CH11357.

## Declarations

All authors have read and approved the manuscript and agree with its submission to MDM. The authors have no conflicts of interest to declare.

