## Supplementary material for "Emulator-based Bayesian calibration of the CISNET colorectal cancer models"

#### Figure S1. Model outputs coverage of targets

##### *S1.1 SimCRC*

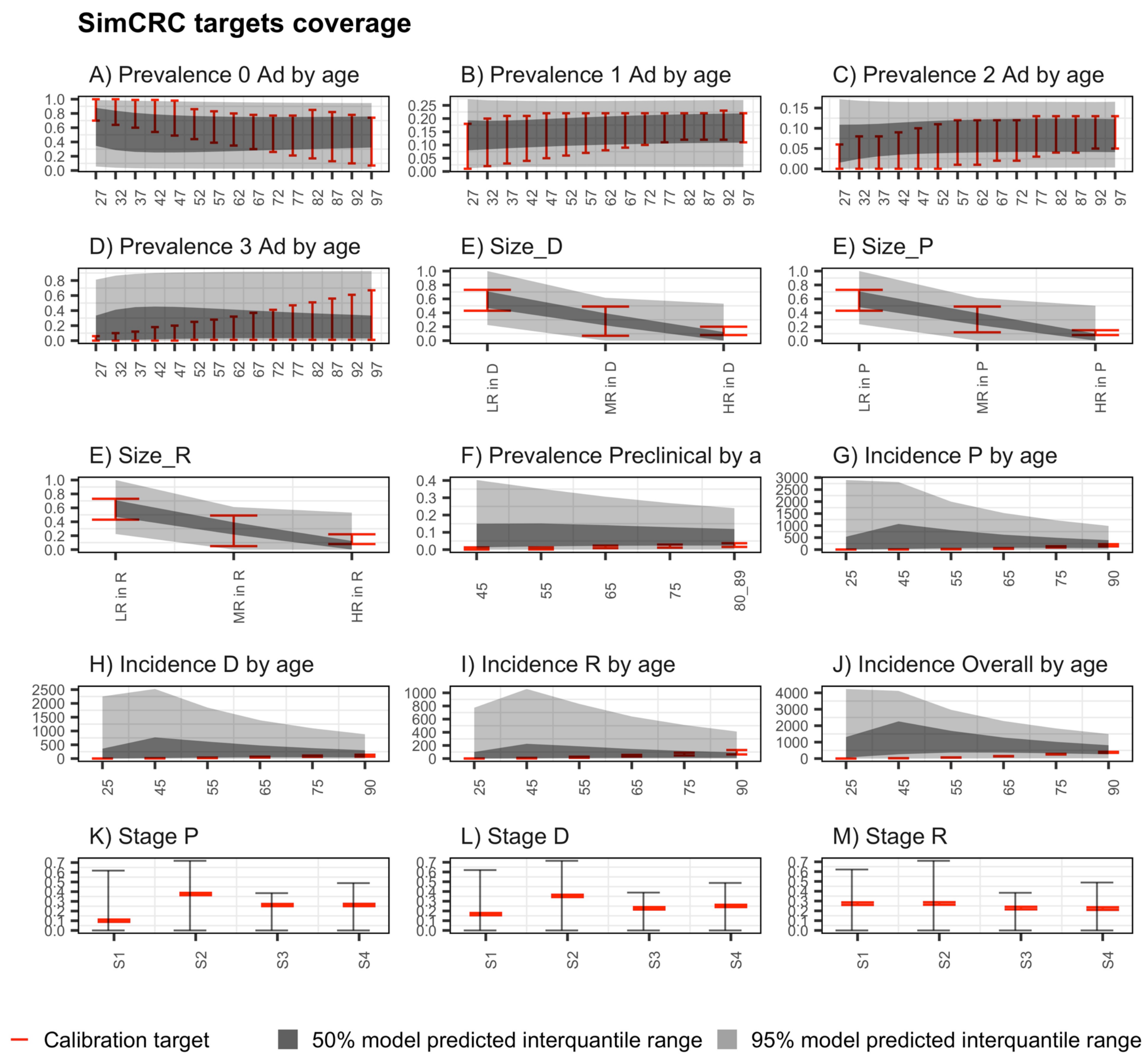

#### *S1.2 MISCAN-Colon*

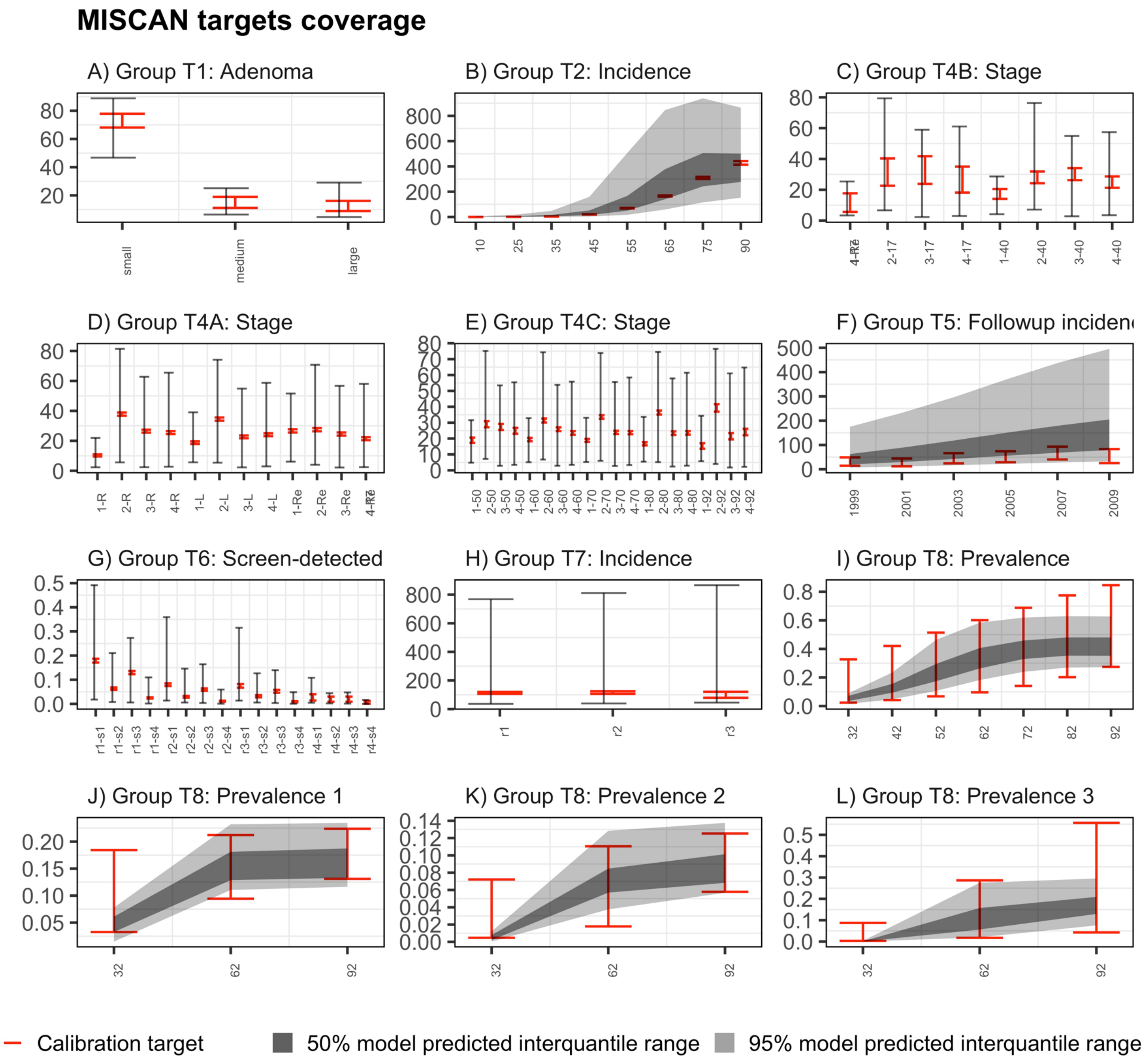

##### *S1.3 CRC-SPIN*

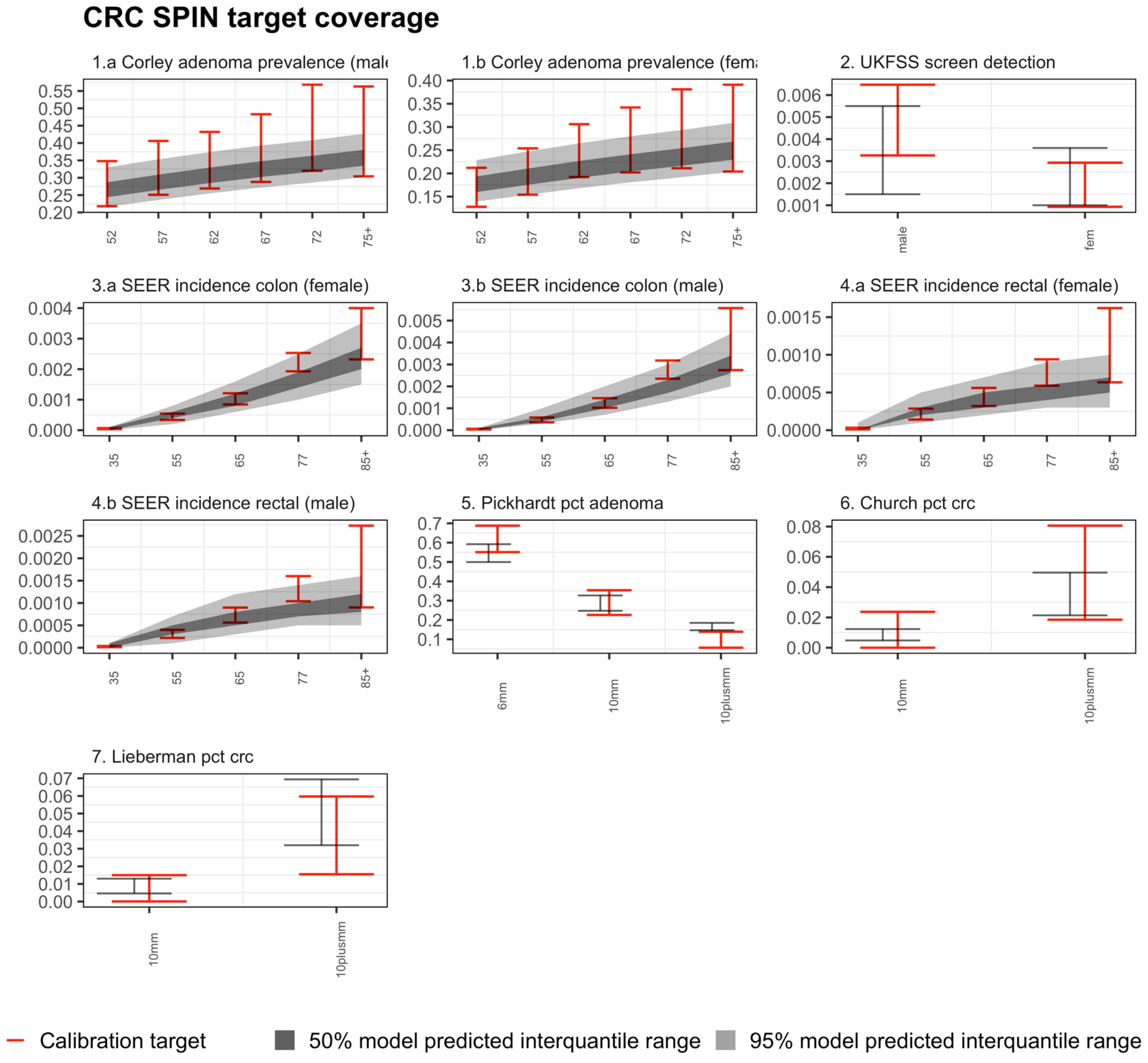

CRC SPIN target coverage
1.a Corley adenoma prevalence (male
1.b Corley adenoma prevalence (fema
2. UKFSS screen detection
3.a SEER incidence colon (female)
3.b SEER incidence colon (male)
4.a SEER incidence rectal (female)
4.b SEER incidence rectal (male)
5. Pickhardt pct adenoma
6. Church pct crc
7. Lieberman pct crc
52
57
62
67
72
75+
male
fem
35
55
65
77
85+
6mm
10mm
10plusmm
Calibration target
50% model predicted interquantile range
95% model predicted interquantile range

**Figure S2. R-hat values for convergence diagnostic and Effective-sample size for efficiency diagnostic.**

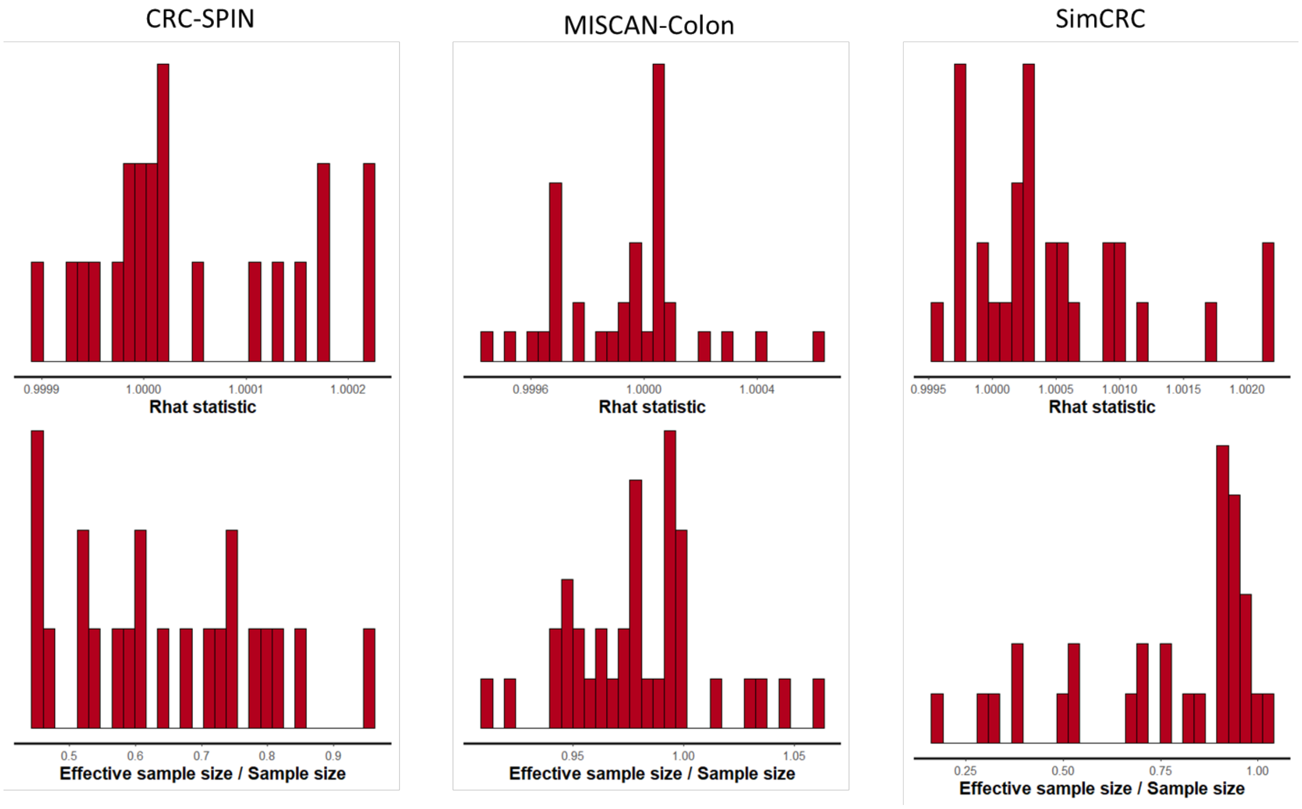

### Figure S3. Prior and posterior distributions of the calibrated input parameters for the three CISNET-CRC models

#### *S3.1 SimCRC*

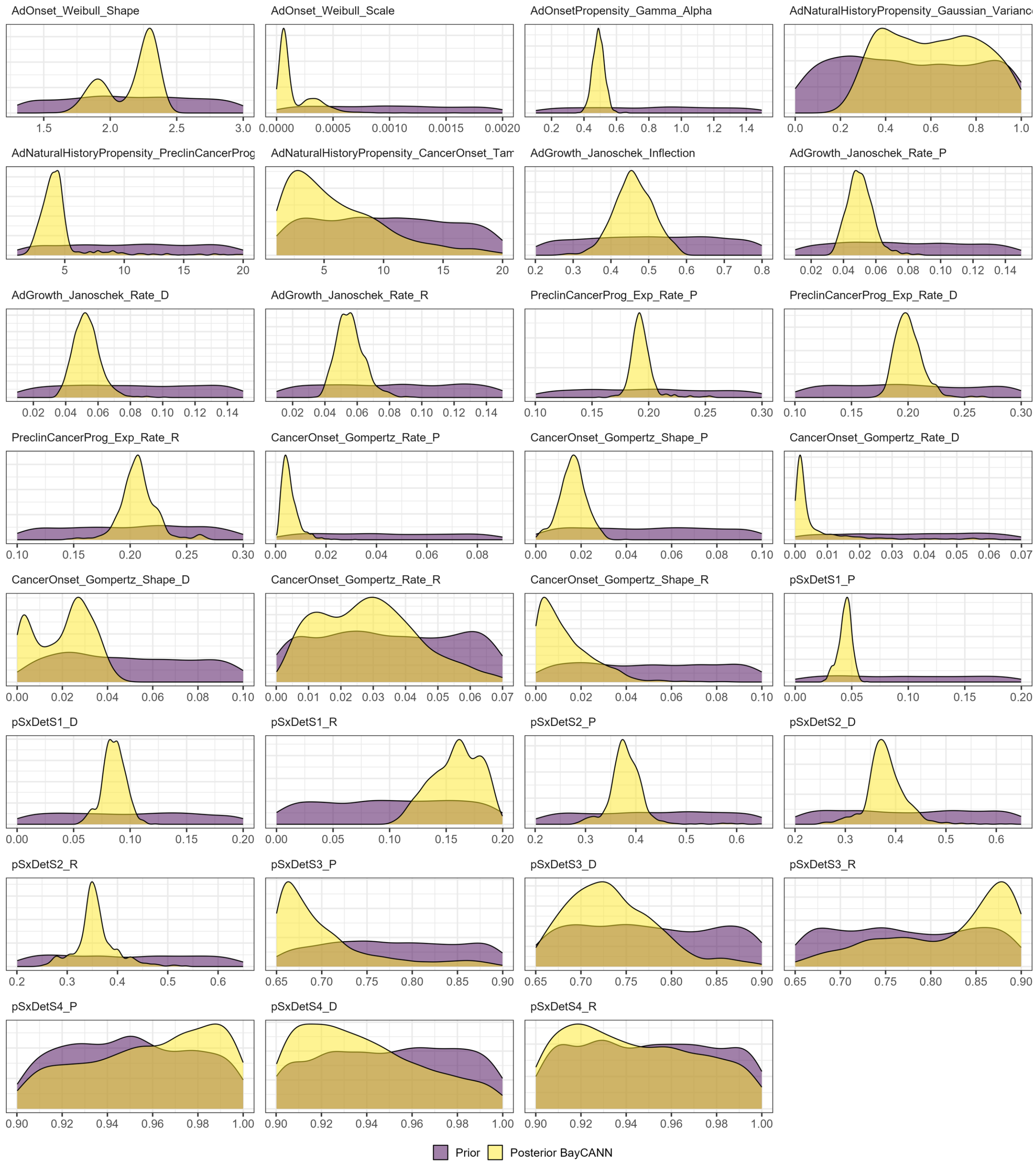

#### *S3.2 MISCAN-Colon*

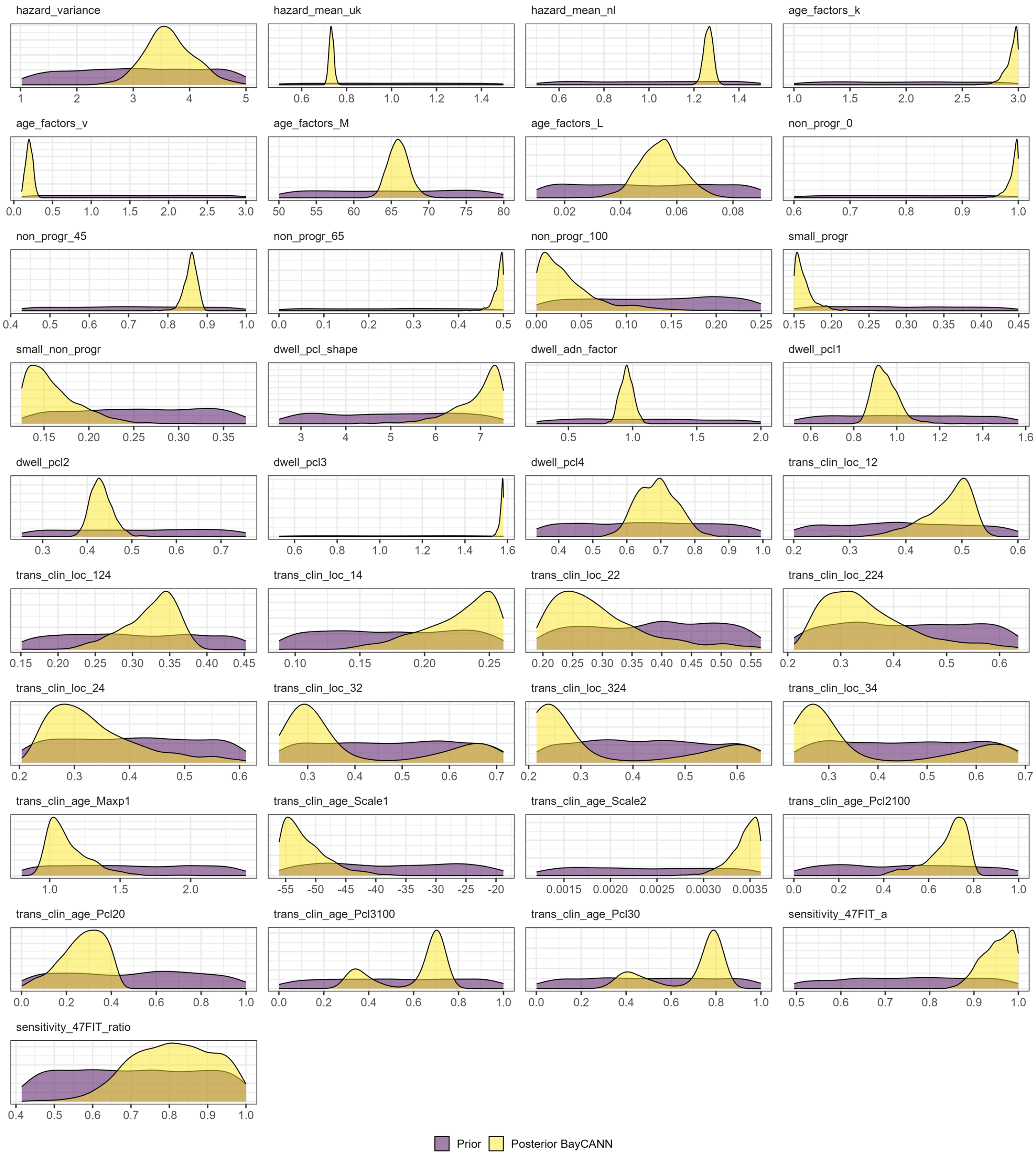

##### *S3.3 CRC-SPIN*

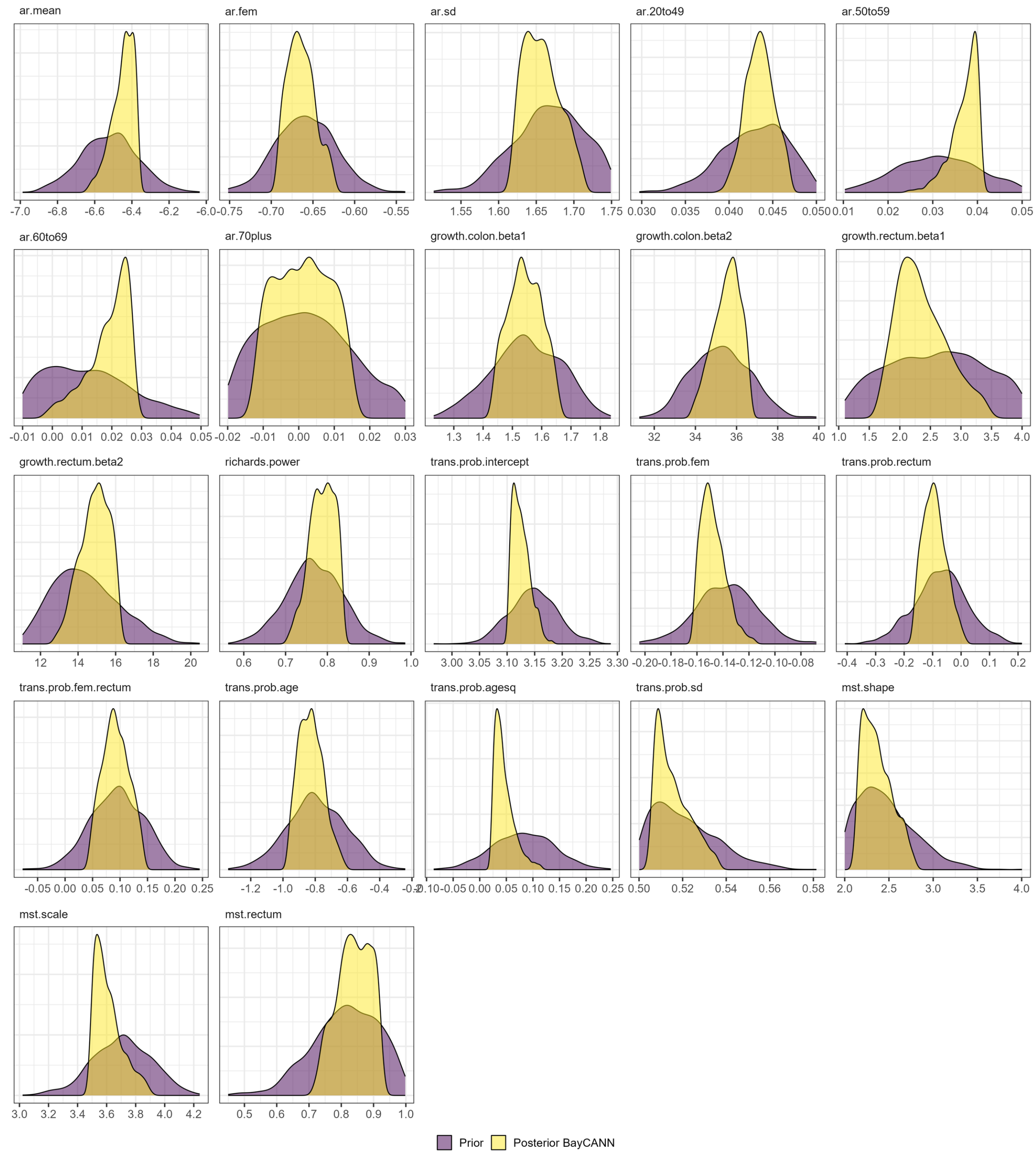


Note: Originally, the prior distributions of CRC-SPIN were set as uniform. However, due to the LHS draws stopping early when predictions fall out of range, this generates a non-uniform prior distribution.

**Figure S4. Joint posterior distribution of the calibrated input parameters for the three CISNET-CRC models.**

#### S4.1 SimCRC

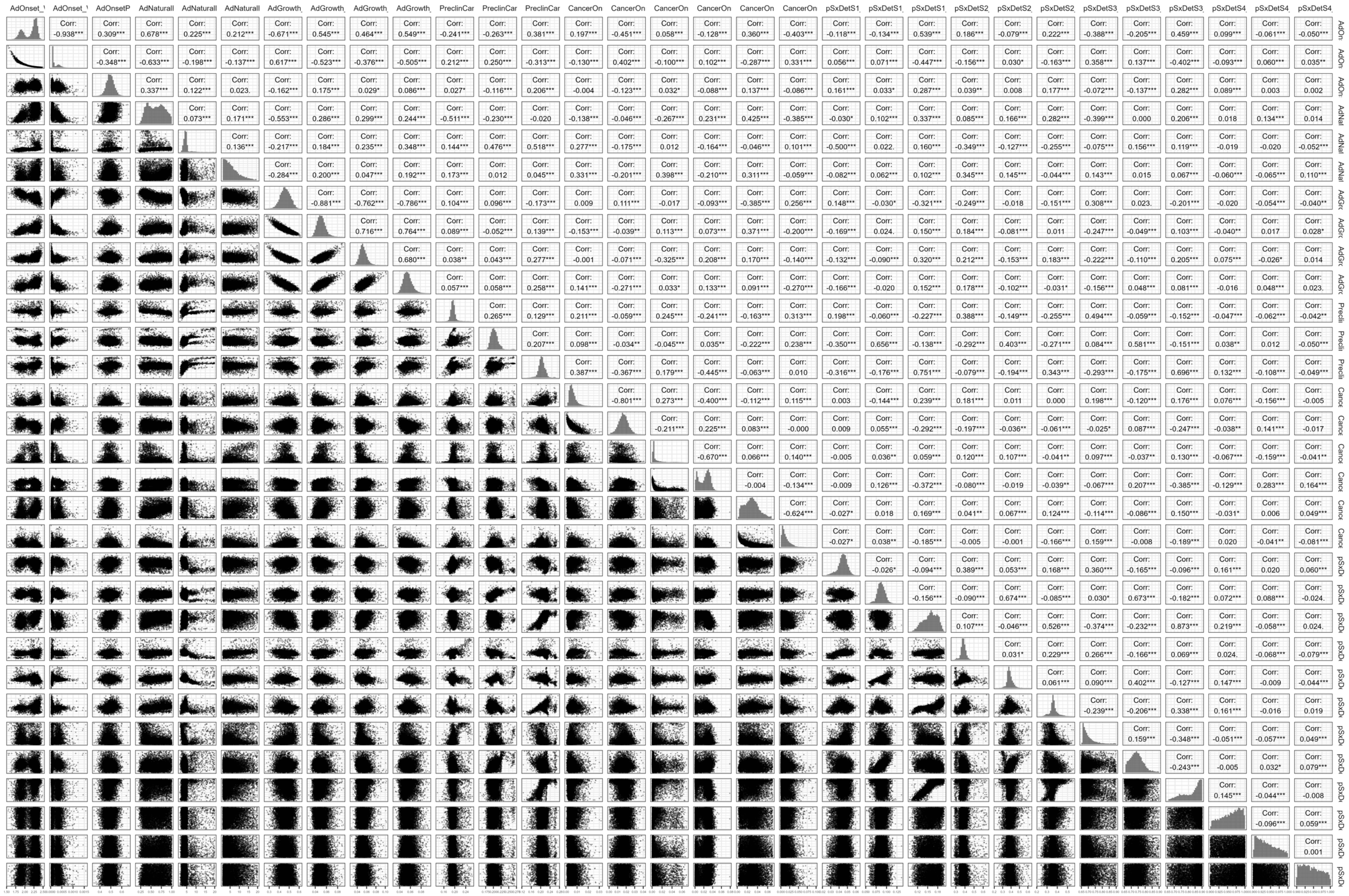

#### *S4.2 MISCAN-Colon*

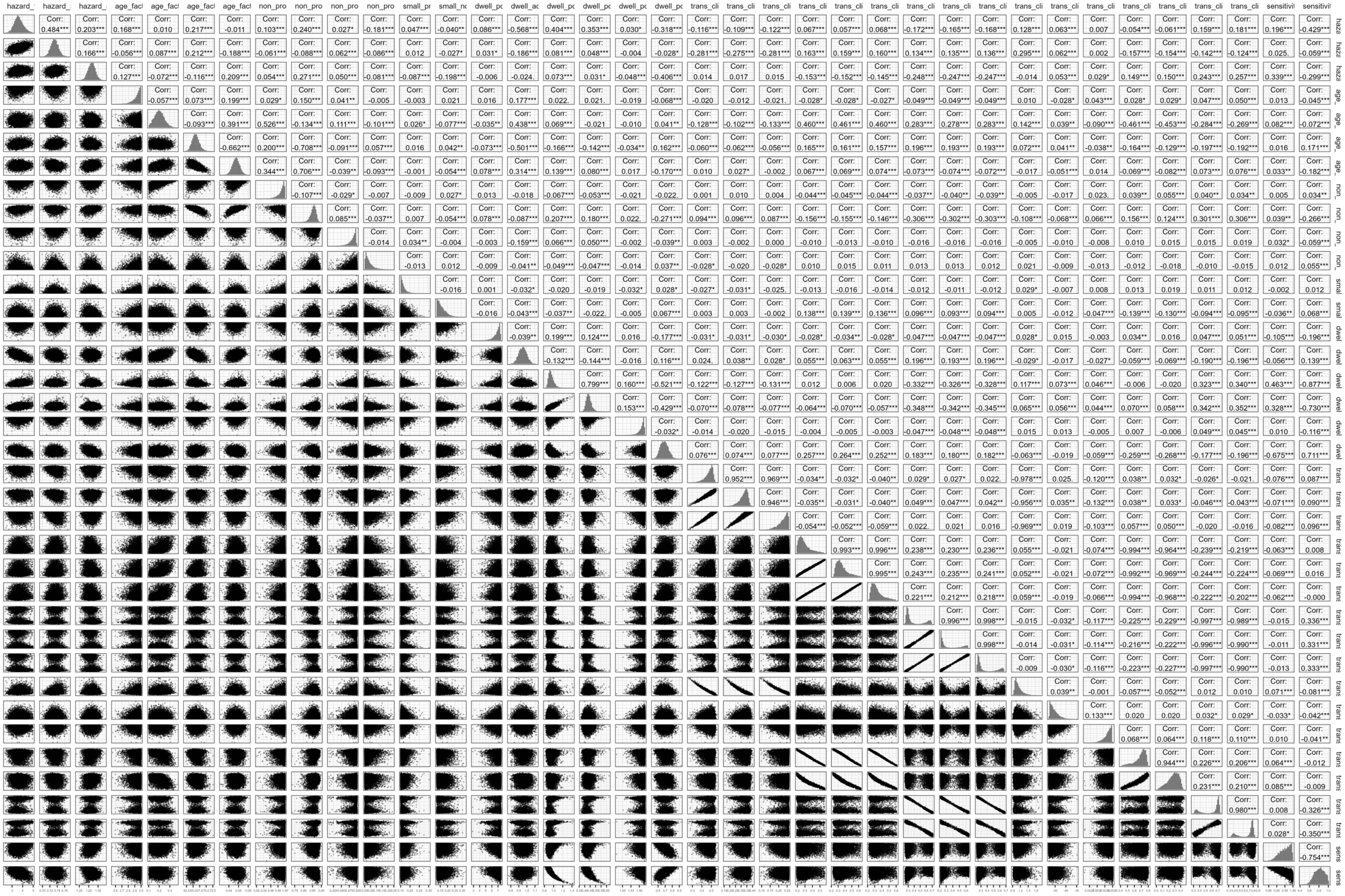

#### *S4.3 CRC-SPIN*

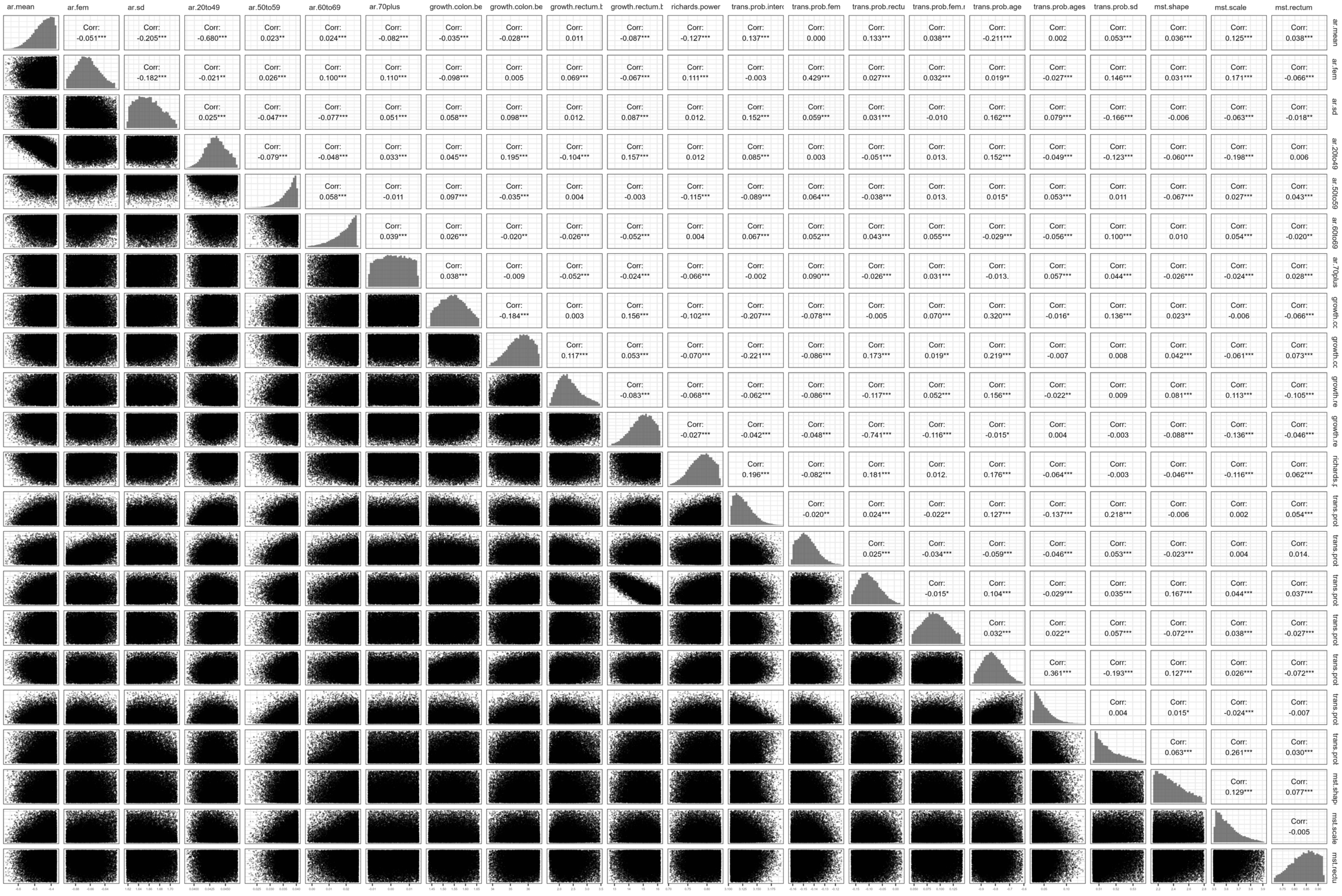
ar.mean
ar.fem
ar.sd
ar.20to49
ar.50to59
ar.60to69
ar.70plus
growth.colon.be
growth.colon.be
growth.rectum.b
growth.rectum.b
richards.power
trans.prob.interc
trans.prob.fem
trans.prob.rectu
trans.prob.fem.r
trans.prob.age
trans.prob.ages
trans.prob.sd
mst.shape
mst.scale
mst.rectum
Corr: -0.051***
Corr: -0.205***
Corr: -0.680***
Corr: 0.023**
Corr: 0.024***
Corr: -0.082***
Corr: -0.035***
Corr: -0.028***
Corr: 0.011
Corr: -0.087***
Corr: -0.127***
Corr: 0.137***
Corr: 0.000
Corr: 0.133***
Corr: 0.038***
Corr: -0.211***
Corr: 0.002
Corr: 0.053***
Corr: 0.036***
Corr: 0.125***
Corr: 0.038***
ar.mean
Corr: -0.182***
Corr: -0.021**
Corr: 0.026***
Corr: 0.100***
Corr: 0.110***
Corr: -0.098***
Corr: 0.005
Corr: 0.069***
Corr: -0.067***
Corr: 0.111***
Corr: -0.003
Corr: 0.429***
Corr: 0.027***
Corr: 0.032***
Corr: 0.019**
Corr: -0.027***
Corr: 0.146***
Corr: 0.031***
Corr: 0.171***
Corr: -0.066***
ar.fem
Corr: 0.025***
Corr: -0.047***
Corr: -0.077***
Corr: 0.051***
Corr: 0.058***
Corr: 0.098***
Corr: 0.012.
Corr: 0.087***
Corr: 0.012.
Corr: 0.152***
Corr: 0.059***
Corr: 0.031***
Corr: -0.010
Corr: 0.162***
Corr: 0.079***
Corr: -0.166***
Corr: -0.006
Corr: -0.063***
Corr: -0.018**
ar.sd
Corr: -0.079***
Corr: -0.048***
Corr: 0.033***
Corr: 0.045***
Corr: 0.195***
Corr: -0.104***
Corr: 0.157***
Corr: 0.012
Corr: 0.085***
Corr: 0.003
Corr: -0.051***
Corr: 0.013.
Corr: 0.152***
Corr: -0.049***
Corr: -0.123***
Corr: -0.060***
Corr: -0.198***
Corr: 0.006
ar.20to49
Corr: 0.058***
Corr: -0.011
Corr: 0.097***
Corr: -0.035***
Corr: 0.004
Corr: -0.003
Corr: -0.115***
Corr: -0.089***
Corr: 0.064***
Corr: -0.038***
Corr: 0.013.
Corr: 0.015*
Corr: 0.053***
Corr: 0.011
Corr: -0.067***
Corr: 0.027***
Corr: 0.043***
ar.50to59
Corr: 0.039***
Corr: 0.026***
Corr: -0.020**
Corr: -0.026***
Corr: -0.052***
Corr: 0.004
Corr: 0.067***
Corr: 0.052***
Corr: 0.043***
Corr: 0.055***
Corr: -0.029***
Corr: -0.056***
Corr: 0.100***
Corr: 0.010
Corr: 0.054***
Corr: -0.020**
ar.60to69
Corr: 0.038***
Corr: -0.009
Corr: -0.052***
Corr: -0.024***
Corr: -0.066***
Corr: -0.002
Corr: 0.090***
Corr: -0.026***
Corr: 0.031***
Corr: -0.013.
Corr: 0.057***
Corr: 0.044***
Corr: -0.026***
Corr: -0.024***
Corr: 0.028***
ar.70plus
Corr: -0.184***
Corr: 0.003
Corr: 0.156***
Corr: -0.102***
Corr: -0.207***
Corr: -0.078***
Corr: -0.005
Corr: 0.070***
Corr: 0.320***
Corr: -0.016*
Corr: 0.136***
Corr: 0.023**
Corr: -0.006
Corr: -0.066***
growth.cc
Corr: 0.117***
Corr: 0.053***
Corr: -0.070***
Corr: -0.221***
Corr: -0.086***
Corr: 0.173***
Corr: 0.019**
Corr: 0.219***
Corr: -0.007
Corr: 0.008
Corr: 0.042***
Corr: -0.061***
Corr: 0.073***
growth.cc
Corr: -0.083***
Corr: -0.068***
Corr: -0.062***
Corr: -0.086***
Corr: -0.117***
Corr: 0.052***
Corr: 0.156***
Corr: -0.022**
Corr: 0.009
Corr: 0.081***
Corr: 0.113***
Corr: -0.105***
growth.re
Corr: -0.027***
Corr: -0.042***
Corr: -0.048***
Corr: -0.741***
Corr: -0.116***
Corr: -0.015*
Corr: 0.004
Corr: -0.003
Corr: -0.088***
Corr: -0.136***
Corr: -0.046***
growth.re
Corr: 0.196***
Corr: -0.082***
Corr: 0.181***
Corr: 0.012.
Corr: 0.176***
Corr: -0.064***
Corr: -0.003
Corr: -0.046***
Corr: -0.116***
Corr: 0.062***
richards.p
Corr: -0.020**
Corr: 0.024***
Corr: -0.022**
Corr: 0.127***
Corr: -0.137***
Corr: 0.218***
Corr: -0.006
Corr: 0.002
Corr: 0.054***
trans.prol
Corr: 0.025***
Corr: -0.034***
Corr: -0.059***
Corr: -0.046***
Corr: 0.053***
Corr: -0.023***
Corr: 0.004
Corr: 0.014.
trans.prol
Corr: -0.015*
Corr: 0.104***
Corr: -0.029***
Corr: 0.035***
Corr: 0.167***
Corr: 0.044***
Corr: 0.037***
trans.prol
Corr: 0.032***
Corr: 0.022**
Corr: 0.057***
Corr: -0.072***
Corr: 0.038***
Corr: -0.027***
trans.prol
Corr: 0.361***
Corr: -0.193***
Corr: 0.127***
Corr: 0.026***
Corr: -0.072***
trans.prol
Corr: 0.004
Corr: 0.015*
Corr: -0.024***
Corr: -0.007
trans.prol
Corr: 0.063***
Corr: 0.261***
Corr: 0.030***
trans.prol
Corr: 0.129***
Corr: 0.077***
mst.shap
Corr: -0.005
mst.scale
mst.rectu

**Figure S5. Internal validation for CRC models.**

***S5.1 SimCRC***

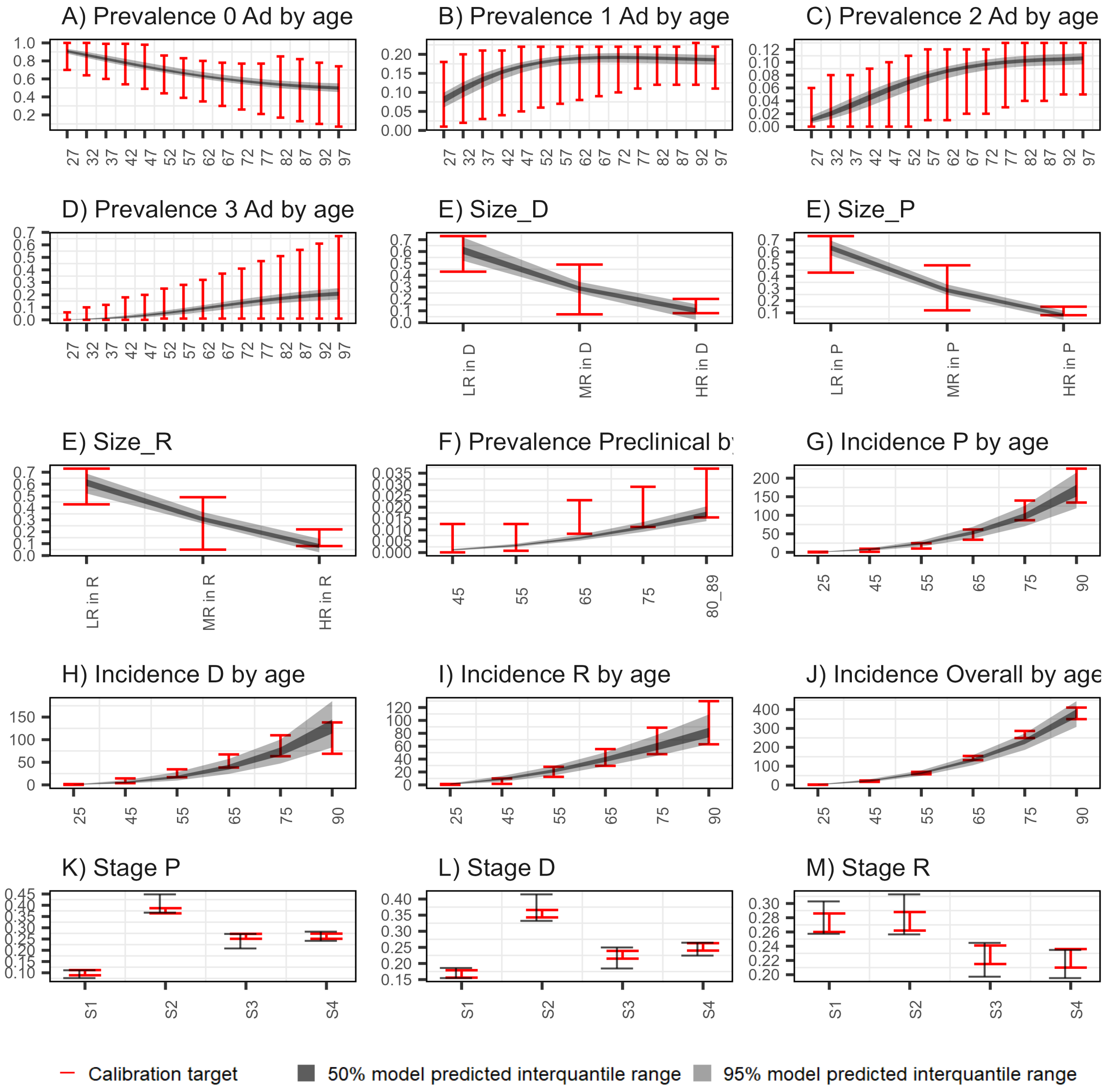

##### *S5.2 MISCAN-Colon*

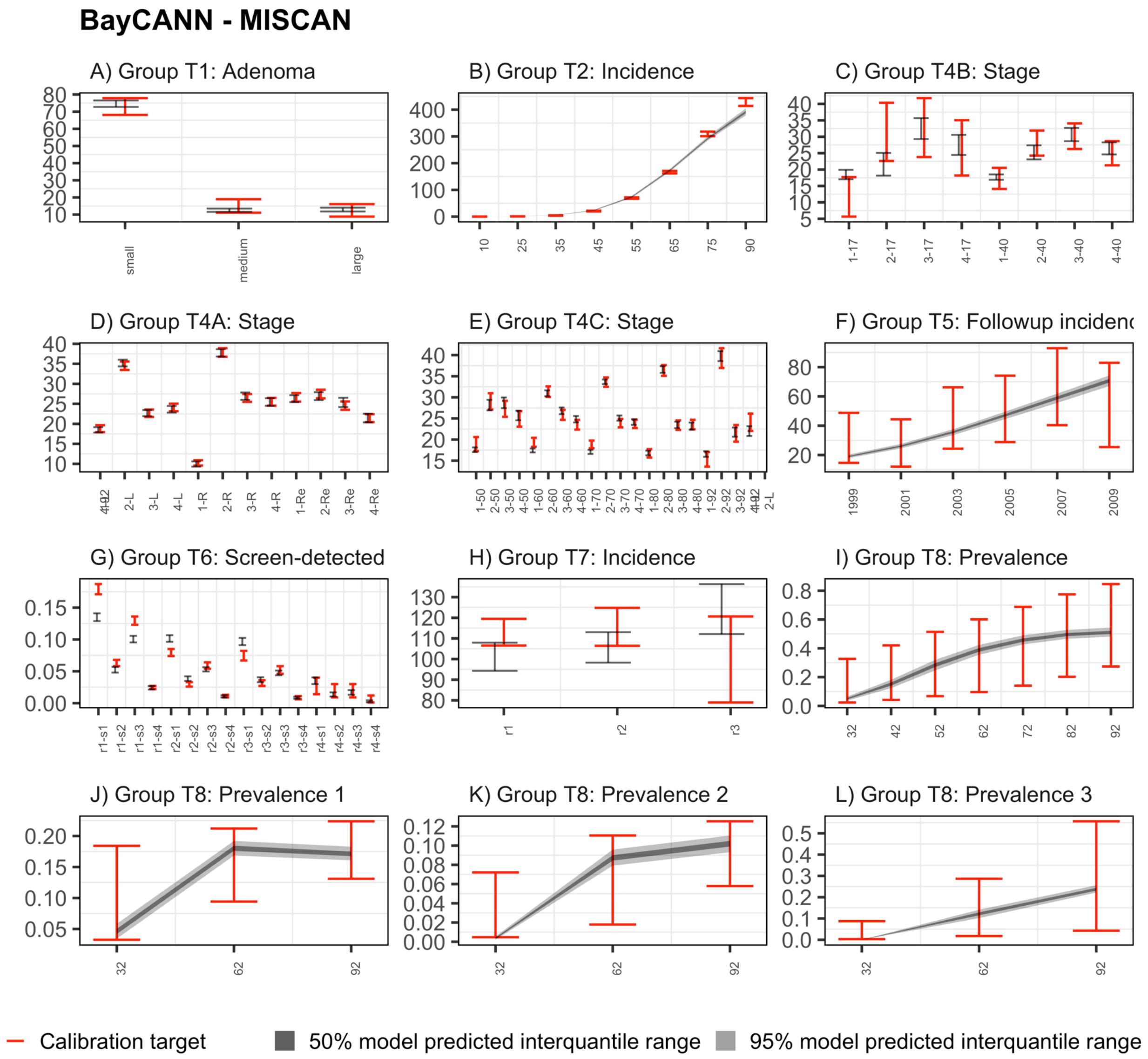

##### *S5.3 CRC-SPIN*

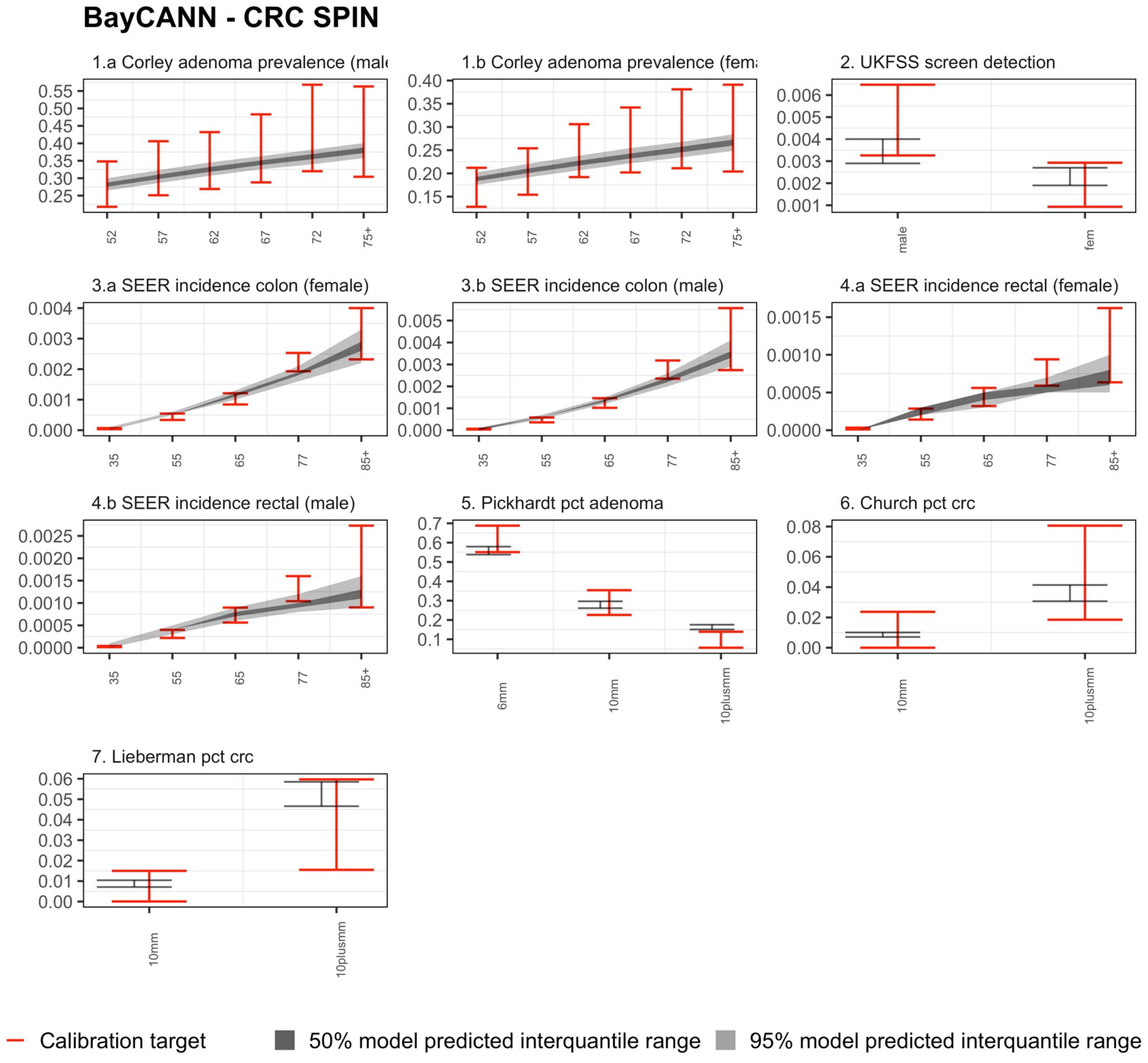
BayCANN - CRC SPIN
1.a Corley adenoma prevalence (male
1.b Corley adenoma prevalence (fema
2. UKFSS screen detection
3.a SEER incidence colon (female)
3.b SEER incidence colon (male)
4.a SEER incidence rectal (female)
4.b SEER incidence rectal (male)
5. Pickhardt pct adenoma
6. Church pct crc
7. Lieberman pct crc
52
57
62
67
72
75+
male
fem
35
55
65
77
85+
6mm
10mm
10plusmm
Calibration target
50% model predicted interquantile range
95% model predicted interquantile range

**Figure S6. Prediction performance of model outputs for the three CRC models ( inputs and outputs are scaled within -1 to 1).**

#### *S6.1 SimCRC*

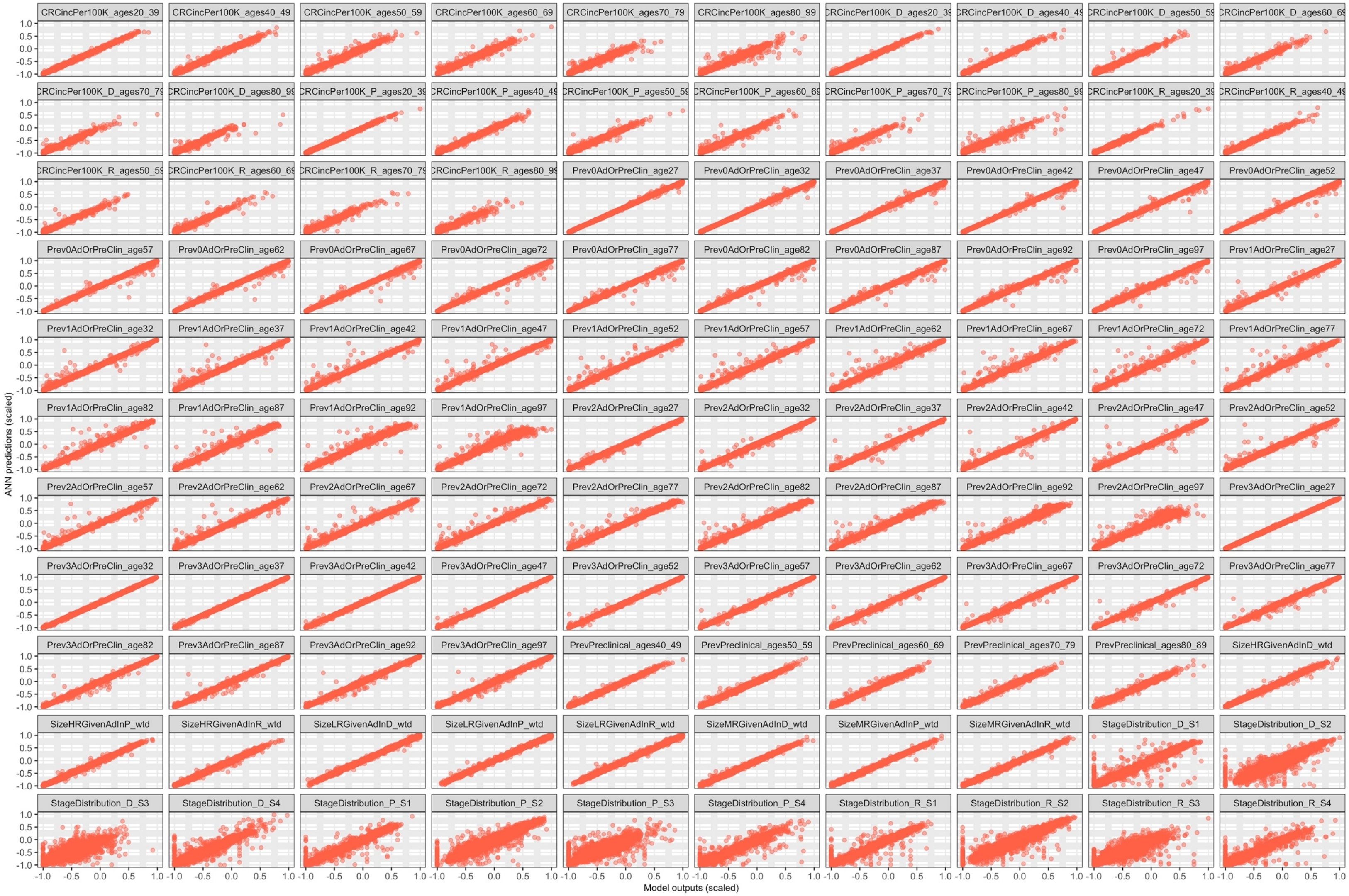

#### *S6.2 MISCAN-Colon*

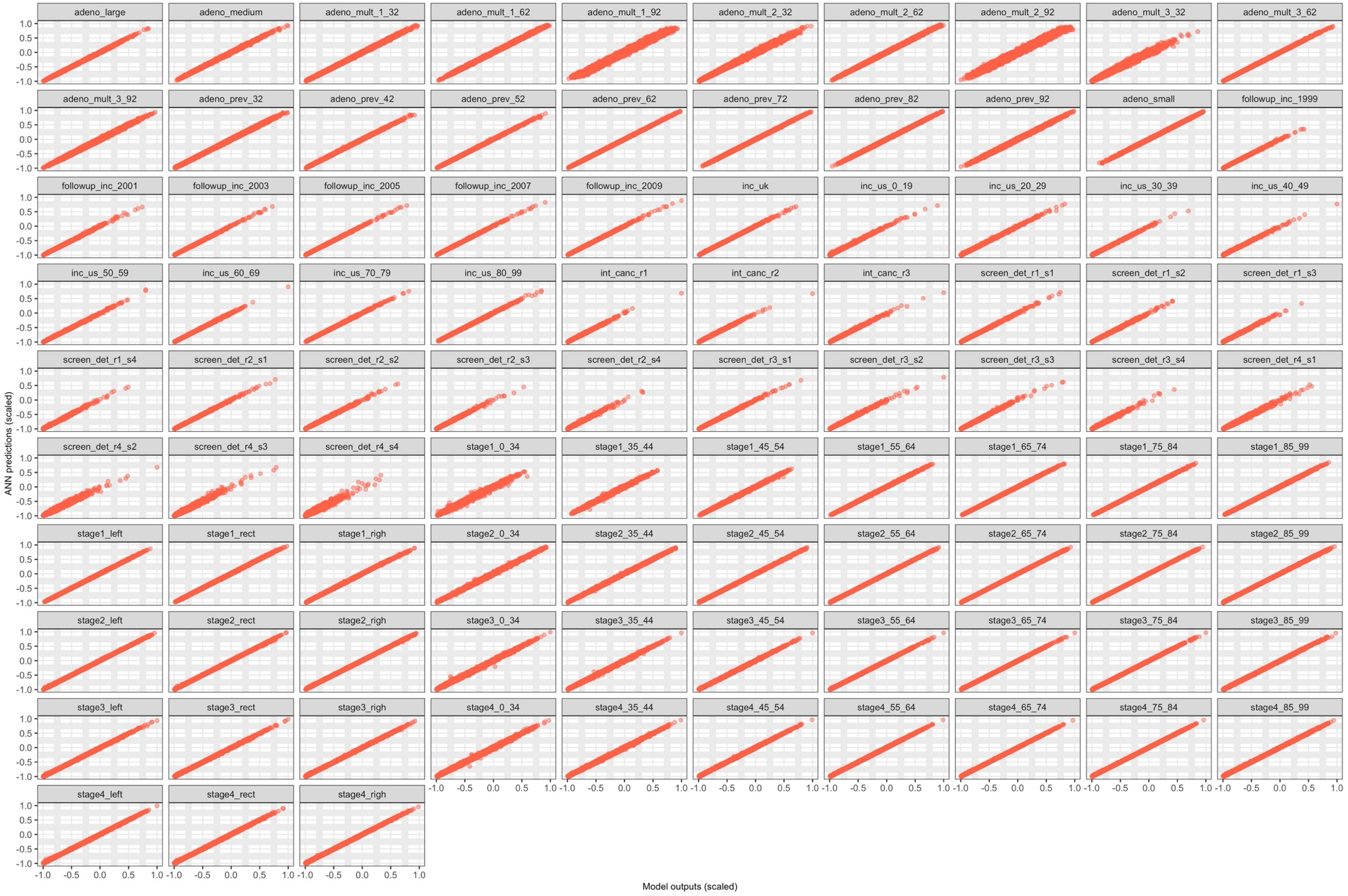

##### *S6.3 CRC-SPIN*

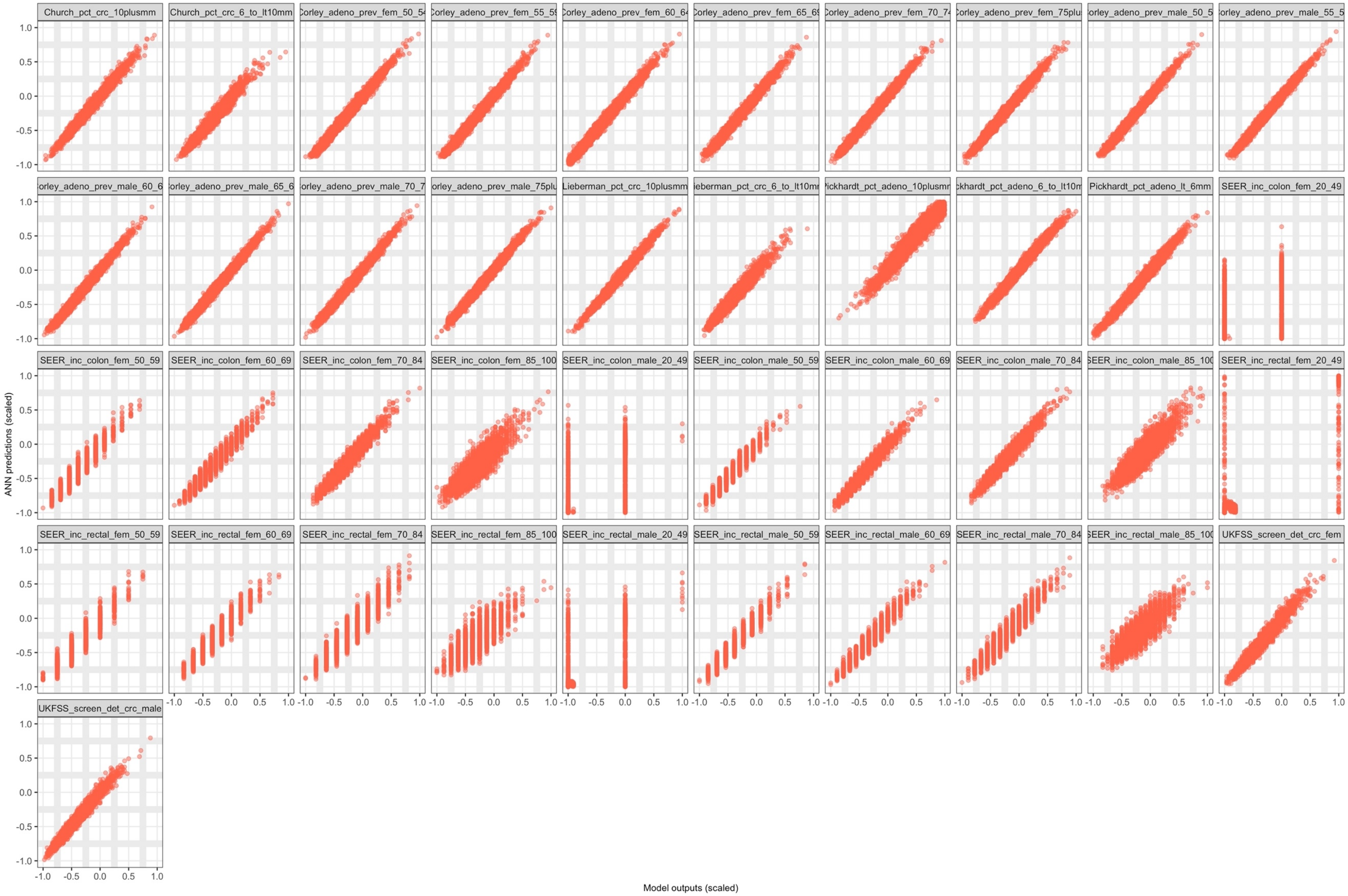


Note: The early stop LHS draws for CRC-SPIN resulted in few outputs in low incidence levels, particularly in the age range of 20-49, as shown in the third and fourth row of the S6.3 figure.

**Figure S7. ANN training time vs. total parameters in the ANN**

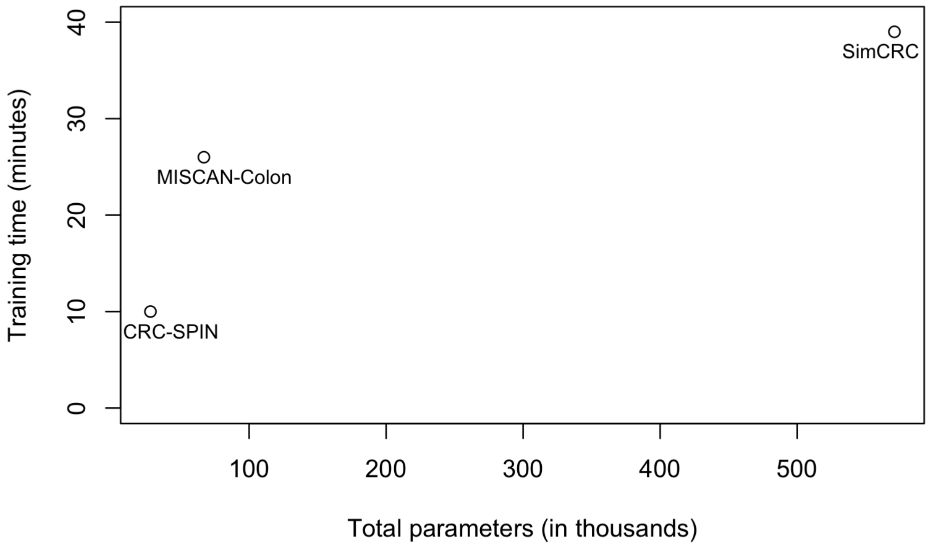


ANN structure and training time

| | Inputs | Hidden layers | Hidden nodes | Outputs | Total ANN trained parameters | Time (minutes) |
|---|---|---|---|---|---|---|
| SimCRC | 30 | 4 | 360 | 110 | 571,070 | 39 |
| MISCAN-Colon | 37 | 4 | 114 | 93 | 67,467 | 26 |
| CRC-SPIN | 22 | 1 | 140 | 41 | 28,741 | 10 |

### Table S8. Calibrated parameters for the three CRC CISNET Models

**Table S8.1 SimCRC**

| Parameter | Description |
|---|---|
| AdOnset_Weibull_Shape | Shape parameter for the risk of developing an adenoma |
| AdOnset_Weibull_Scale | Scale parameter for the risk of developing an adenoma |
| AdOnset_PersonRisk_Gamma_Alpha | Individual risk parameter |
| AdGrowthMult_Variance | Variance for adenoma growth function |
| AdGrowthMult_ScaleTerm | Scale for adenoma growth function |
| Janoschek_Inflection | Inflection parameter for adenoma growth function |
| Janoschek_Rate_P | Rate parameter for growth function for an adenoma in the proximal colon |
| Janoschek_Rate_D | Rate parameter for growth function for an adenoma in the distal colon |
| Janoschek_Rate_R | Rate parameter for growth function for an adenoma in the rectum |
| CancerOnset_Weibull_Shape_P | Shape parameter for cancer onset function for an adenoma in the proximal colon |
| CancerOnset_Weibull_Shape_D | Shape parameter for cancer onset function for an adenoma in the distal colon |
| CancerOnset_Weibull_Shape_R | Shape parameter for cancer onset function for an adenoma in the rectum |
| CancerOnset_Weibull_Scale_P | Scale parameter for cancer onset function for an adenoma in the proximal colon |
| CancerOnset_Weibull_Scale_D | Scale parameter for cancer onset function for an adenoma in the distal colon |
| CancerOnset_Weibull_Scale_R | Scale parameter for cancer onset function for an adenoma in the rectum |
| Exp_Rate_PreclinProg_P | Rate parameter for progression of a preclinical cancer in the proximal colon |
| Exp_Rate_PreclinProg_D | Rate parameter for progression of a preclinical cancer in the distal colon |
| Exp_Rate_PreclinProg_R | Rate parameter for progression of a preclinical cancer in the rectum |
| pSxDetS1_P | Probability of detecting a stage I proximal colon cancer by symptoms |
| pSxDetS1_D | Probability of detecting a stage I distal colon cancer by symptoms |
| pSxDetS1_R | Probability of detecting a stage I rectal cancer by symptoms |
| pSxDetS2_P | Probability of detecting a stage II proximal colon cancer by symptoms |
| pSxDetS2_D | Probability of detecting a stage II distal colon cancer by symptoms |
| pSxDetS2_R | Probability of detecting a stage II rectal cancer by symptoms |
| pSxDetS3_P | Probability of detecting a stage III proximal colon cancer by symptoms |
| pSxDetS3_D | Probability of detecting a stage III distal colon cancer by symptoms |
| pSxDetS3_R | Probability of detecting a stage III rectal cancer by symptoms |
| pSxDetS4_P | Probability of detecting a stage IV proximal colon cancer by symptoms |
| pSxDetS4_D | Probability of detecting a stage IV distal colon cancer by symptoms |
| pSxDetS4_R | Probability of detecting a stage IV rectal cancer by symptoms |

**Table S8.2 MISCAN-Colon**

| Parameter | description |
|---|---|
| hazard_variance | The variance of the Gamma distribution. |
| hazard_mean_uk | Relative risk in the UK compared to the US. |
| hazard_mean_nl | Relative risk in the Netherlands compared to the US. |
| age_factors_k | The value of the horizontal asymptote of the generalized logistic function. |
| age_factors_logb | |
| age_factors_logv | A measure for the slope at the point of inflection of the generalized logistic function and the line symmetry in the point of inflection. |
| age_factors_M | The location of the point of inflection (x-coordinate). |
| non_progr_0 | The probability that an adenoma is non-progressive at age 0 |
| non_progr_45 | The probability that an adenoma is non-progressive at age 45 |
| non_progr_65 | The probability that an adenoma is non-progressive at age 65 |
| non_progr_100 | The probability that an adenoma is non-progressive at age 100 |
| small_progr | The probability that a progressive adenoma does not grow larger than 10mm before it becomes a preclinical cancer |
| small_non_progr | The probability that a non-progressive adenoma never grows larger than 10mm. |
| dwell_pcl_shape | Determines the shape of the Weibull distribution for all preclinical cancer states. |
| dwell_adn_factor | The ratios are multiplied with this parameter to determine the mean dwell time for each adenoma state. |
| dwell_pcl1 | Scale of the Weibull distribution for the preclinical cancer state 1 |
| dwell_pcl2 | Scale of the Weibull distribution for the preclinical cancer state 2 |
| dwell_pcl3 | Scale of the Weibull distribution for the preclinical cancer state 3 |
| dwell_pcl4 | Scale of the Weibull distribution for the preclinical cancer state 4 |
| trans_clin_loc_12 | The probability that a preclinical cancer starts showing clinical symptoms in stage 1 for rectal cancers. |
| trans_clin_loc_124 | The probability that a preclinical cancer starts showing clinical symptoms in stage 2 for cancers in the left colon. |
| trans_clin_loc_14 | The probability that a preclinical cancer starts showing clinical symptoms in stage 1 for cancers in the right colon |
| trans_clin_loc_22 | The probability that a preclinical cancer starts showing clinical symptoms in stage 2 for rectal cancers. |
| trans_clin_loc_224 | The probability that a preclinical cancer starts showing clinical symptoms in stage 2 for cancers in the left colon. |
| trans_clin_loc_24 | The probability that a preclinical cancer starts showing clinical symptoms in stage 2 for cancers in the right colon |
| trans_clin_loc_32 | The probability that a preclinical cancer starts showing clinical symptoms in stage 3 for rectal cancers. |
| trans_clin_loc_324 | The probability that a preclinical cancer starts showing clinical symptoms in stage 3 for cancers in the left colon. |
| trans_clin_loc_34 | The probability that a preclinical cancer starts showing clinical symptoms in stage 3 for cancers in the right colon |
| trans_clin_age_Maxp1 | For stage 1 cancers, the transition probability is represented by quadratic function of age |
| trans_clin_age_Scale1 | Scale factor 1 for transition probability from stage 1 |

| | |
|---|---|
| trans_clin_age_Scale2 | Scale factor 2 for transition probability from stage 1 |
| trans_clin_age_Pcl2100 | Transition probabilities to from stage 2 clinical at age 100 |
| trans_clin_age_Pcl20 | Transition probabilities to from stage 2 clinical at age 0 |
| trans_clin_age_Pcl3100 | Transition probabilities from stage 3 to clinical at age 100 |
| trans_clin_age_Pcl30 | Transition probabilities from stage 3 to clinical at age 0 |
| sensitivity_47FIT_a | The sensitivity of the FIT short before clinical diagnosis |
| sensitivity_47FIT_ratio | The sensitivity of the FIT long before clinical diagnosis |

**Table S8.3 CRC-SPIN**

| Parameter | description |
|---|---|
| ar.mean | Baseline log-risk |
| ar.sd | Standard dev., baseline log-risk |
| ar.fem | Female |
| ar.20to49 | Age effect |
| ar.50to59 | Age effect |
| ar.60to69 | Age effect |
| ar.70plus | Age effect |
| growth.colon.beta1 | Shape, colon |
| growth.colon.beta2 | Scale, colon |
| growth.rectum.beta1 | Shape, rectum |
| growth.rectum.beta2 | Scale, rectum |
| richards.power | Richard's power |
| trans.prob.intercept | Intercept |
| trans.prob.fem | Female (vs male) |
| trans.prob.rectum | Rectal (vs colon) |
| trans.prob.fem.rectum | Female and Rectal |
| trans.prob.age | Age at initiation |
| trans.prob.agesq | Age at initiation sq. |
| trans.prob.sd | Standard dev., Trans. Prob. |
| mst.scale | Scale |
| mst.shape | Shape |
| mst.rectum | Rectal (vs colon) |
